# Atlas of functional connectivity relationships across rare and common genetic variants, traits, and psychiatric conditions

**DOI:** 10.1101/2021.05.21.21257604

**Authors:** Clara A. Moreau, Kuldeep Kumar, Annabelle Harvey, Guillaume Huguet, Sebastian Urchs, Elise A. Douard, Laura M. Schultz, Hanad Sharmarke, Khadije Jizi, Charles-Olivier Martin, Nadine Younis, Petra Tamer, Thomas Rolland, Jean-Louis Martineau, Pierre Orban, David Shin, Ana Isabel Silva, Jeremy Hall, Marianne B.M. van den Bree, Michael J. Owen, David E. J. Linden, Aurelie Labbe, Anne M. Maillard, Tomasz J. Nowakowski, Sarah Lippé, Carrie E. Bearden, Laura Almasy, David C. Glahn, Paul M. Thompson, Thomas Bourgeron, Pierre Bellec, Sebastien Jacquemont

## Abstract

Polygenicity and pleiotropy are key properties of the genomic architecture of psychiatric disorders. An optimistic interpretation of polygenicity is that genomic variants converge on a limited set of mechanisms at some level from genes to behavior. Alternatively, convergence may be minimal or absent.

We took advantage of brain connectivity, measured by resting-state functional MRI (rs-fMRI), as well as rare and common genomic variants to understand the effects of polygenicity and pleiotropy on large-scale brain networks, a distal step from genes to behavior. We processed ten rs-fMRI datasets including 32,988 individuals, to examine connectome-wide effects of 16 copy number variants (CNVs), 10 polygenic scores, 6 cognitive and brain morphometry traits, and 4 idiopathic psychiatric conditions.

Although effect sizes of CNVs on connectivity were correlated to cognition and number of genes, increasing polygenicity was associated with decreasing effect sizes on connectivity. Accordingly, the effect sizes of polygenic scores on connectivity were 6-fold lower compared to CNVs. Despite this heterogeneity of connectivity profiles, multivariate analysis identified convergence of genetic risks and psychiatric disorders on the thalamus and the somatomotor network. Based on spatial correlations with transcriptomic data, we hypothesize that excitatory thalamic neurons may be primary contributors to brain alteration profiles shared across genetic risks and conditions. Finally, pleiotropy measured by genetic and transcriptomic correlations between 38 pairs of conditions/traits showed significant concordance with connectomic correlations, suggesting a substantial causal genetic component for shared connectivity.

Such findings open avenues to delineate general mechanisms - amenable to intervention - across conditions and genetic risks.

**One sentence summary:** Effects of rare and common genomic variants on brain functional connectivity shed light on the impact of polygenicity and pleiotropy in psychiatry.

## Introduction

Genetic pleiotropy and polygenicity are key features of psychiatric conditions (*1*). Common and rare variants both contribute to risk. Genetic correlation (rG) is a measure of genetic overlap between conditions, also referred to as pleiotropy. It is moderate to high between schizophrenia (SZ), bipolar disorder (BIP), and major depressive disorder (MDD) and milder between these three conditions and autism spectrum disorder (ASD) (*2–4*). Varying levels of genetic correlations are also observed between conditions, brain morphometry, cognitive and behavioral traits (*5, 6*). Although overlap has mostly been computed for common variants (single nucleotide polymorphisms, SNPs), data shows that pleiotropy also applies to rare variants such as copy number variants (CNVs)(*7, 8*), which are associated with a broad range of cognitive phenotypes and overlapping psychiatric diagnoses.

Polygenicity is particularly problematic for studying risk and mechanisms. The latter has been modeled for SNPs, using polygenic scores (PGS), which are weighted sums of genetic risk alleles (*9*). Similar models applied to rare variants show that the effects of CNVs on cognition are the sum of individual effects of genes encompassed in the CNV, weighted by their sensitivity to gene dosage (*10, 11*).

An optimistic interpretation of polygenicity is that genomic variants may converge on a smaller set of mechanisms -amenable to intervention- at some level from gene transcription to microcircuits to large-scale connectivity networks to behavior (*12*). A less optimistic interpretation embraces complexity. In this scenario, convergence would occur very late -or not at all- in the ‘pathway’ to behavior. Polygenicity would therefore lead to “poly- connectivity”; that is, each genomic variant leads to a mostly distinct large-scale brain connectivity profile (**Figure 1A**)(*13*). In the latter scenario, individuals with the same diagnosis would present a broad array of distinct alterations. It is unknown whether polygenicity leads to shared or distinct alterations of brain connectivity pathways implicated in psychiatric conditions.

**Figure 1.**
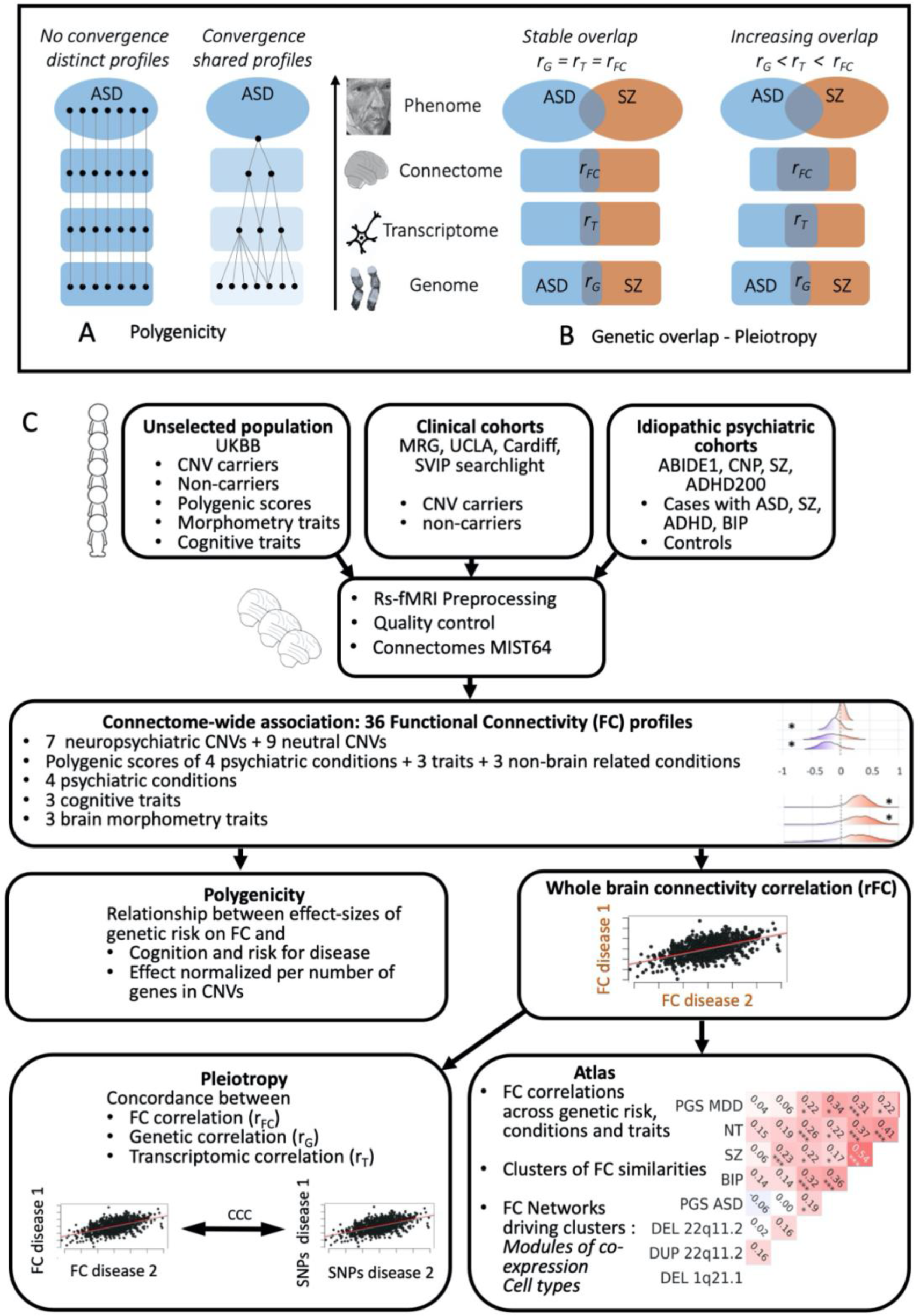
Schematic of key hypotheses and methods flowchart. Legend: a) Left side: scenario with little convergence and extreme diversity (pessimistic scenario): 1000 genes are associated with 1000 mainly distinct transcriptomic, brain, and behavioral profiles of ASD. Right side: optimistic scenario; important mechanistic convergence from genes to behavior with alterations of FC profiles shared across genetic variants, resulting in a limited number of mechanistic subtypes. b)Left side: levels of pleiotropy are stable from gene to functional connectivity profiles. Right side: Pleiotropy increases from genes to functional connectivity. c) Methods flowchart. Abbreviations: CNV: copy number variation; FC: functional connectivity; ASD: Autism Spectrum Disorder; SZ: schizophrenia; CNP: Consortium for Neuropsychiatric Phenomics, MRG: Montreal rare genomic disorder.

Understanding the effects of polygenicity and pleiotropy on brain architecture may therefore help to identify and target broad psychiatric risk pathways amenable to common therapeutic interventions (*12*).

The organization of large-scale functional networks in the brain can be inferred using resting-state functional MRI (rs-fMRI). This imaging technique measures spontaneous, low-frequency temporal synchronization of the activity in different brain regions during rest (*14, 15*). Several studies demonstrated that functional networks are related to the spatial distribution of gene expression in the brain, especially genes linked to ion channel activity and synaptic function (*16, 17*). Functional connectivity (FC) has gained traction, characterizing increasingly reproducible patterns of FC alterations associated with psychiatric conditions (*18*). Studies have provided critical insight into the architecture of brain networks involved in neuropsychiatric disorders and have demonstrated overlap at the connectivity level (rFC) between conditions (*19*) It is unknown whether brain FC overlap between conditions reflects genetic pleiotropy.

### Knowledge gap

How polygenicity and pleiotropy reflect aspects of large-scale brain functional networks in neuropsychiatric disorders is unknown.

Genetics-first studies investigate individuals selected on the basis of specific genomic variants irrespective of psychiatric symptoms or diagnoses. Such approaches offer opportunities to investigate effects of genetic factors on FC irrespective of clinical diagnoses. Deletions and duplications at the 16p11.2 and 22q11.2 loci have been associated with mirror effects on global FC (i.e., average connectivity strength across all regions in the brain)(*20*). In contrast to diagnostic-first studies, effect-sizes for high-risk neuropsychiatric mutations were of similar magnitude for neuroimaging features and behavioral traits (*19*). Connectivity profiles of the thalamus, somatomotor, posterior insula and cingulate showed substantial similarities between neuropsychiatric CNVs and idiopathic ASD and SZ, but not attention deficit hyperactivity disorder (ADHD). Beyond these two genomic loci, nothing is known about the effects of rare variants on network connectivity. As well, little is known about the FC effects of PGS (additive common genomic risk) for psychiatric conditions (*21*). Overall, the relationship between FC and neuropsychiatric variants is understudied.

### Our overarching aim was to investigate brain connectivity relationships between multiple genetic risks, traits, and diseases to provide insight into mechanisms underlying polygenicity and pleiotropy in psychiatric conditions

Specifically, we aimed to: 1) Characterize the FC profiles of rare neuropsychiatric CNVs ranging from mono- or oligo-genic CNVs (i.e., involving a single gene or small gene sets) to large polygenic CNVs (eg. n=50 genes for 22q11.2 deletion) and PGS; 2) Investigate the relationship between the level of polygenicity and effects of genomic variants on FC; 3) Investigate the relationship of previously established genetic correlations (referred as pleiotropy) to FC similarities between conditions and traits; 4) Identify brain networks associated with genetic risks, psychiatric diseases, and traits.

To this end, we processed all rs-fMRI data using the same pipeline in n=32,988 individuals. We performed 36 connectome-wide association studies (CWAS, **Figure 1B**) on 1) 1,003 carriers of one among 7 neuropsychiatric and 9 non-psychiatric CNVs identified from 4 clinical cohorts and the UK Biobank; 2) 30,185 and 174 non-CNV carriers from the UK Biobank and clinical cohorts respectively, 3) 778 individuals with idiopathic ASD, SZ, BIP or ADHD from 4 datasets and their respective 848 controls (**Table 1**). The following data have never been published: 876 CNV carriers, all PGS, all brain morphometry traits, and two out of the four genetics-first cohorts.

**Table 1.**
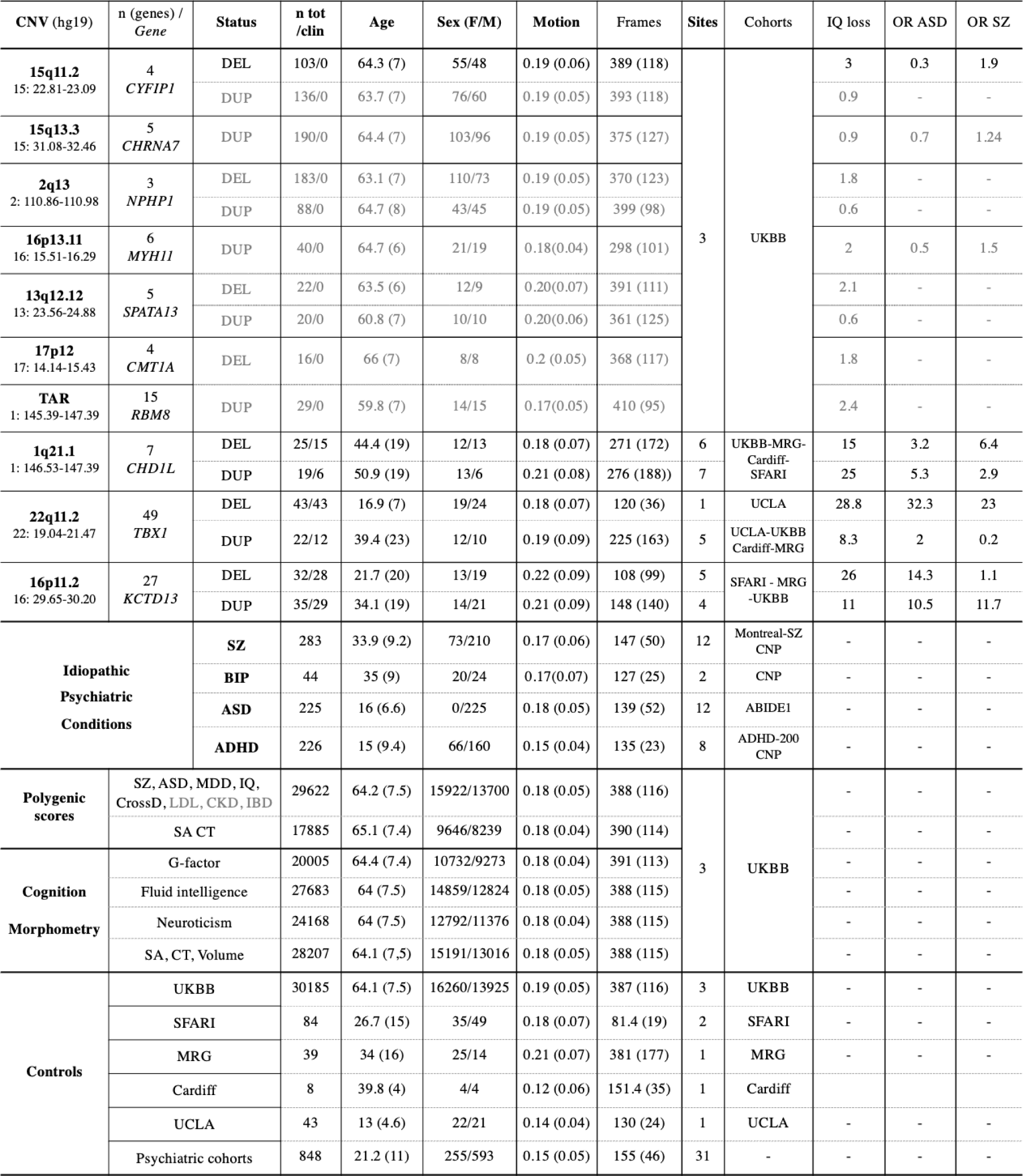
Data demographics. Legend: CNV carriers, individuals with idiopathic psychiatric conditions, and controls after MRI quality control. Chr: chromosome number, coordinates are presented in Megabases (Mb, Hg19). n = tot /Clin: total number of participants /number of participants clinically ascertained. Age (in years); M: male; Motion: framewise displacement (in mm). Quantitative variables are expressed as the mean ± standard deviation. All sites scanned controls and sensitivity analyses were performed to investigate the potential bias introduced by differences in scanning site, age, and sex. IQ loss: mean decrease in IQ points associated with each CNV (10, 11). Odd-ratios (OR) for the enrichment of CNVs in ASD and schizophrenia were previously published (25–27, 67–72). OR for the enrichment of CNVs in ADHD were not available. The 9 non-psychiatric CNVs (light grey font) were defined as variants without any previous association with psychiatric conditions in large cases control studies (24–26, 41), and detailed information relative to diagnosis, IQ, and motion, are available in Supplementary results. The 3 non-brain-related PGS are in a light grey font. We excluded connectomes of subjects scanned before 2018 (n=11,331) for PGS-SA and PGS-CT because they were used in the discovery GWAS. Abbreviations: DEL: deletion; DUP: duplication; SZ: schizophrenia, ASD: Autism Spectrum Disorder; ADHD: Attention-Deficit / Hyperactivity-Disorder, BIP: Bipolar disorder, MDD: Major Depression Disorder, CrossD: Cross-disorder, LDL: Low-Density Lipoprotein, IBD: Inflammatory Bowel Disease, CKD: Chronic Kidney Disease; SA: Surface Area, CT: Cortical Thickness, CNP: Consortium for Neuropsychiatric Phenomics, MRG: Montreal rare genomic disorder; IQ: intelligence quotient.

## Results

### Effect sizes of 36 FC profiles across genetic risk, conditions, cognitive and brain morphometry traits

We computed brain-wide FC profiles with and without global signal adjustment (GSA; **Figures 2A-E, Supplementary table 4**). FC-profiles were defined as the 2,080 β values of 2,080 connections -connectivity between 64 functional parcels- obtained from the contrast of cases vs. controls. All connectivity values were z-scored, based on the variance of corresponding control groups. Compared to controls, six CNVs, ASD, SZ, and all of the brain and cognitive traits showed a mild shift in mean FC (mean of all beta values; **Table 2**, **Figure 2A-E**). After GSA, FC profiles remained mutually correlated. It was previously demonstrated that GSA-FC profiles show stronger correlations with cognition(*22*), and reduce confounding effects in multi-site studies (*23*). We, therefore, used GSA-FC profiles for the remainder of this study.

**Figure 2.**
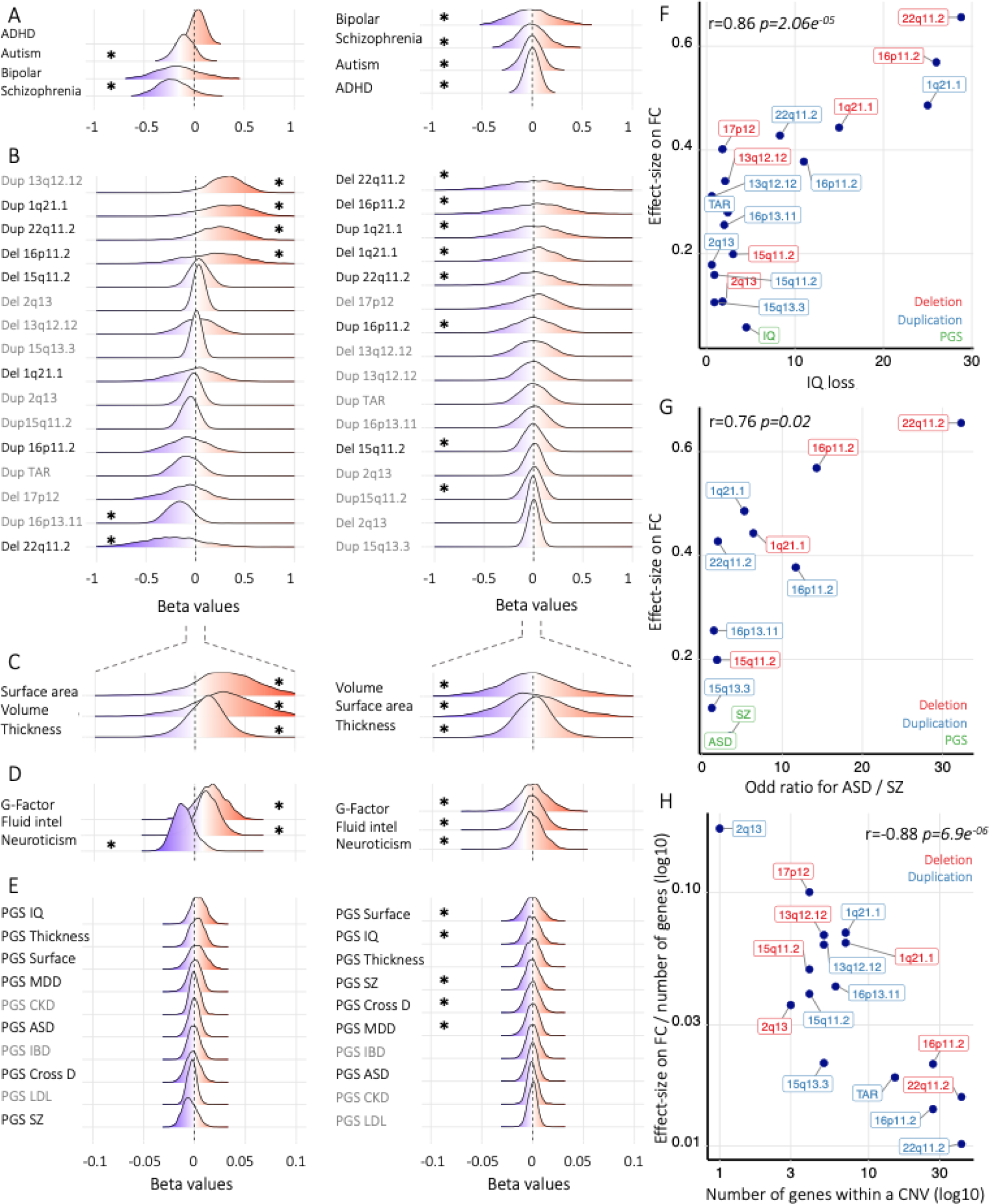
Effect sizes across idiopathic conditions, CNVs, PGS, and traits. Legend: (A-E) Density plots represent the distribution of the 2,080 beta values of each FC-profile obtained by connectome-wide association study (CWAS) for (A) idiopathic condition, (B) CNVs, (C-D) traits, (E) Polygenic scores (PGS). X-axis values of all density plots represent beta values, which were obtained from linear models computed using z-scored connectomes based on the variance of the control group. Left column of density plots shows effects on global FC signal without GSA and stars represent significant shifts of mean connectivity. Right column of density plots shows effect sizes after GSA and stars represent FC-profiles with at least one altered connection surviving FDR. (F) Correlation between previously published effect-sizes of CNVs on IQ (10) and their effect-sizes on FC. The X-axis is the mean decrease in IQ associated with each CNV. (G) Correlation between previously published effect-sizes of CNVs on ASD-SZ risk (24–27) and their effect-sizes on FC. The X-axis for is the Odd ratio of each CNV computed in previously published case-control studies. PGS are not included in the correlation. (H) Correlation between CNV gene content (X-axis) and effect-sizes on FC normalized by the number of genes encompassed in CNVs. Y-axes of all figures (F-H) are effect sizes on FC after GSA (the top decile of beta-values). Abbreviations: ASD: autism spectrum disorder, SZ: schizophrenia, MDD: major depressive disorder, NT: neuroticism, Del: deletion, Dup: duplication, Fluid intel: fluid intelligence, IQ: intelligence quotient, surface area, ADHD: Attention-Deficit / Hyperactivity-Disorder, IBD: Inflammatory bowel disease, LDL: Low-Density Lipoprotein; CKD: Chronic Kidney Disease.

**Table 2.**
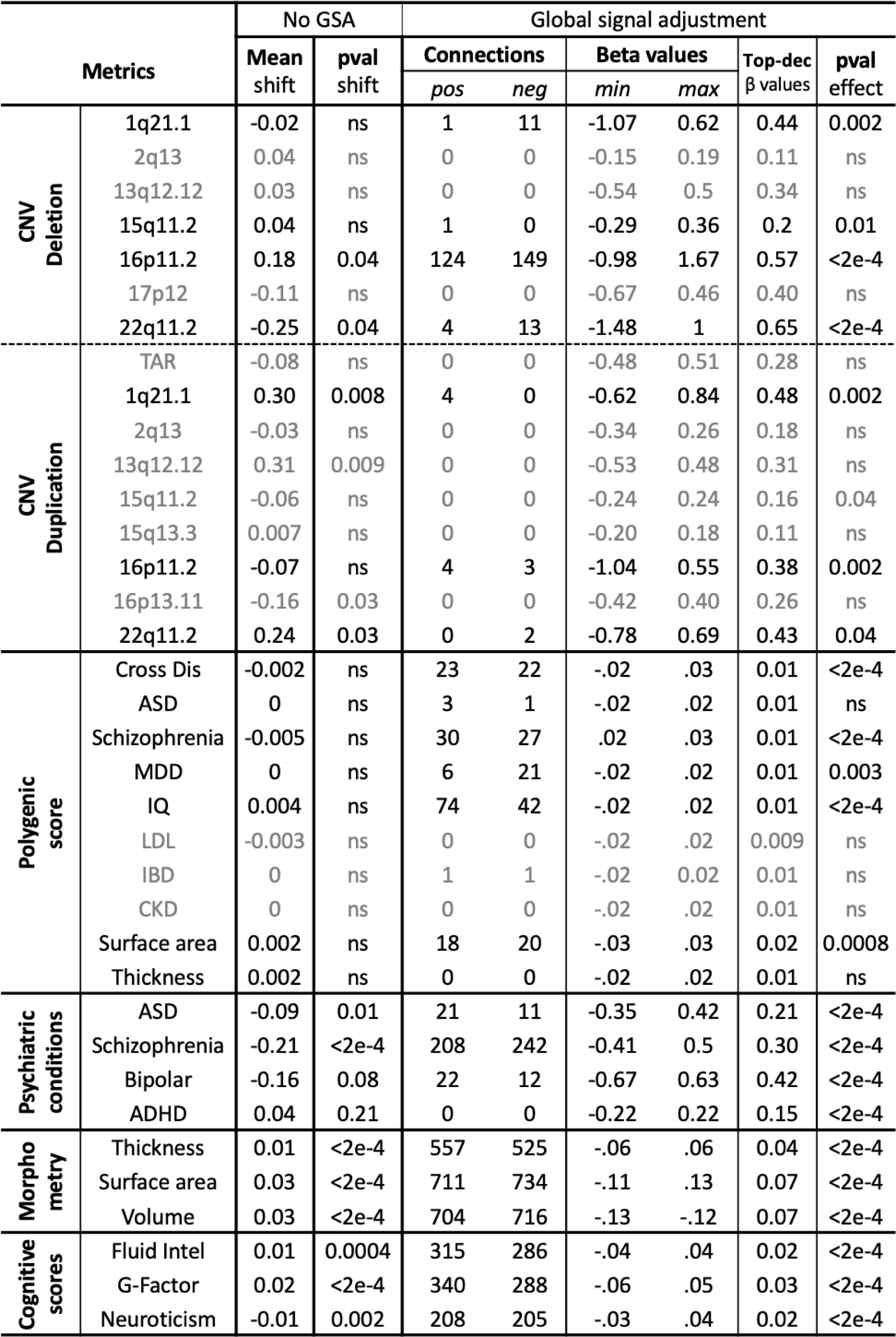
CWAS summary. Legend: The number of significantly altered connections (FDR corrected) for each connectome-wide association study (n=36) after global signal adjustment (GSA) (Supplementary Table 4). DEL: deletion; DUP: duplication; ASD: autism spectrum disorder; SZ: schizophrenia; ADHD: attention deficit hyperactivity disorder. min-max: minimum-maximum of z-scored beta values; top decile: top decile of beta values; Connection pos: number of positive connections surviving FDR; Connection neg: number of negative connections surviving. Abbreviations: DEL: deletion; DUP: duplication; SZ: schizophrenia, ASD: Autism Spectrum Disorder; ADHD: Attention-Deficit / Hyperactivity-Disorder, BIP: Bipolar disorder, MDD: Major Depression Disorder, CrossD: Cross-disorder, LDL: Low-Density Lipoprotein, IBD: Inflammatory Bowel Disease, CKD: Chronic Kidney Disease; SA: Surface Area, CT: Cortical Thickness, IQ: intelligence quotient.

Effect-sizes were largest for neuropsychiatric CNVs followed by psychiatric conditions, brain morphometry, cognitive traits, and PGS. All 7 neuropsychiatric and one out of nine “non-psychiatric” CNVs altered connections that survived false discovery rate (FDR) (2,080 connections, *q*<0.05, **Table 2**). Sensitivity analyses - performing contrasts in 5,000 randomly sampled groups - found the same level of significance compared to FDR procedure (**Table 2**). The 16p11.2 deletion and duplication showed large effects on FC-profiles, which were correlated (*r*=0.7 and 0.83) to the previously published profiles that were based on smaller samples. The previously published 22q11.2 deletion FC profile also showed large effects. 1q21.1 deletion and duplication FC profiles showed moderate to large effects on FC. 15q11.2 deletion and duplications showed the mildest effects among the neuropsychiatric CNVs.

None of the brain-related PGS computed in the UKBB showed mean shifts in global connectivity, but all of them (except cortical thickness and ASD) altered GSA-FC profiles with 5 to 116 connections surviving FDR. The non-brain PGS traits showed no significant effects surviving FDR and the permutation test (**Table 1**, **Figure 2**). Effect sizes were on average one order of magnitude smaller than those observed for CNVs. The correlation between the FC profiles of PGS and their corresponding conditions and traits (rFC) ranged from 0.23 to 0.73 (*p*=0.01 to 0.0001). This suggests that connectivity alterations observed in psychiatric conditions are also observed, to some extent, in healthy individuals at increased risk.

Patients diagnosed with idiopathic SZ, BIP, ASD but not ADHD significantly altered FC compared to controls. For SZ, ADHD, and ASD, profiles were previously published (*20*) but we recomputed them for ADHD and SZ after adding individuals. Correlations between new and previously published profiles were 0.70 and 0.95, respectively.

All cognitive and brain morphometry traits were associated with FC profiles of mild effect-sizes. Sensitivity analyses showed that results were robust to the effects of sex, pooled or matched controls, clinical or non-clinical ascertainment, and medication (Supplemental results).

### Increasing polygenicity decreases functional connectivity signals

Previous studies demonstrated that the effect-size of CNVs on IQ increases approximately linearly with the number of encompassed genes. The number of deleted genes (weighted by dosage sensitivity) predicts IQ with 78% accuracy (*10, 11*). We observed a correlation between the effect size of CNVs on FC and their previously estimated effect size on cognitive ability(*10*) (*r*=0.86, *p*=2.06e^-05^) and risk for either ASD(*24, 25*) or SZ (*26, 27*) (*r*=0.76, *p*=0.02). These relationships appeared non-linear (**Figure 2F, G**), suggesting that effect size on FC may not be a mere additive effect of individual genes encompassed in CNVs. Accordingly, the effect size of deletions and duplications normalized by the number of genes they encompass significantly decreased from a single gene to large polygenic CNVs (**Figure 2H**). This relationship was linear on a log-log scale (*r*=-0.85, *p*=3.0e^-05^). In other words, large multigenic CNVs have much smaller effects on FC than expected based on the number of genes they encompass (additive model). This same decrease in effect-size was observed when accounting for the level and number of dosage-sensitive genes within CNVs (*r*= -0.88, *p*= 6.9e^-06^).

In line with this observation, effect sizes of PGS for ASD, SZ, and IQ (common variants) were approximately 6-fold smaller (**Figure 2F-G**) than those observed for CNVs with similar effect sizes on cognition and risk for ASD and SZ.

### Pleiotropy: Brain connectivity mirrors genetic correlations between conditions and traits

Genetic overlap (measured by SNP-based genetic correlations: rG) is a key feature of the architecture of psychiatric conditions, brain morphometry, and cognitive and behavioral traits (*2, 5, 6*). A similar overlap between conditions has also been shown at the gene expression level (transcriptomic correlations: rT,, **Figure 3A**)(*28*). We asked if FC correlations (rFC) between conditions and traits show similar relationships. Specifically, genetic pleiotropy and polygenicity may imply that genes converge on shared FC alterations leading to an increasing overlap (correlation values) from genes to transcriptome to connectome (**Figure 1AB**).

We first demonstrated a significant concordance between rT and rFC across 10 pairs of conditions and traits (CCC=0.76, 95%CI: [0.41; 0.91] without any bias (bias correction factor = 0.99; **Figure 3B**)).

**Figure 3.**
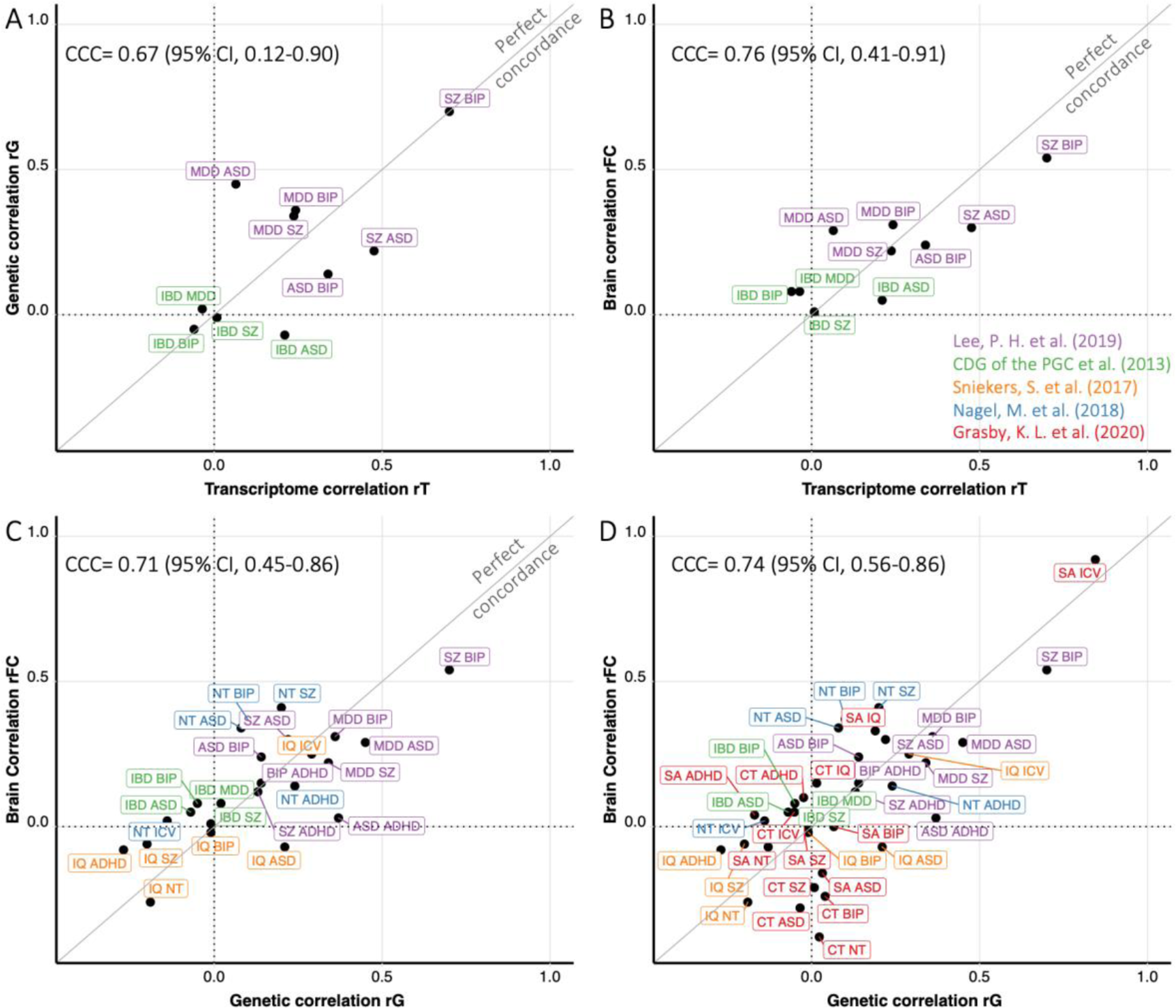
Concordance across genetic, transcriptomic and connectomic correlations. Legend: A) Concordance between genetic correlation and transcriptomic correlation across pairs of conditions and traits (as previously published(28) with updated genetic correlations) b) The same concordance analysis was performed between FC correlations and transcriptomic correlations using the same conditions and traits. c) Concordance between genetic correlation and FC correlations across pairs of conditions and cognitive-behavioral traits. d) The same concordance as is (c) with brain morphometry traits. X and Y axis: r values of correlations. The brain correlations (rFC) represent the correlation between the FC profiles of a pair of conditions-traits. The diagonal represents a perfect concordance. Transcriptomic correlations (28) and genetic correlation values were previously published (Supplementary Table 5). All the correlations values are available in Supplemental Table 5. Colors indicate papers that computed rG : (purple (2), green (73), orange (6), blue (61), red (5)). Since we could not compute the FC profile of MDD and IBD we used the FC profile of their respective PGS. Abbreviations: CCC: concordance correlation coefficient, CI: confidence interval, ASD: autism spectrum disorder, SZ: schizophrenia, BIP: bipolar disorder, MDD: major depressive disorder, NT: neuroticism, Del: deletion, Dup: duplication, Fluid intel: fluid intelligence, SA: surface area, CT: cortical thickness, IQ: intelligence quotient, surface area, ICV: intracranial volume, ADHD: Attention-Deficit / Hyperactivity-Disorder, IBD: Inflammatory bowel disease.

We also showed a significant concordance between rG and rFC across 24 pairs of conditions and traits (CCC=0.71, 95%CI: [0.45; 0.86], bias correction factor = 0.99, **Figure 3C**). In other words, FC overlap between conditions and traits was neither higher nor lower than rG (**Figure 1A**). In a sensitivity analysis, we replaced FC profiles of traits and conditions with the FC profiles of their corresponding PGS. This resulted in the same concordance (CCC=0.71, 95%CI: [0.54;0.83], bias correction factor = 0.97). Finally, adding 3 brain morphometry traits (n=38 pairs) did not change the level of concordance (**Figure 3D**).

### Most networks are affected by genetic risk and conditions

All functional brain networks were affected by neuropsychiatric CNVs, PGS, psychiatric conditions, and traits (**Figure 4**). Basal ganglia-thalamus and somatomotor networks showed over-connectivity across most genetic risk measures and conditions. In contrast, ventral attention and auditory/posterior insula networks were predominantly under-connected. Dysconnectivity in the default mode and limbic networks exhibited the largest effect-sizes (**Figure 4**).

**Figure 4.**
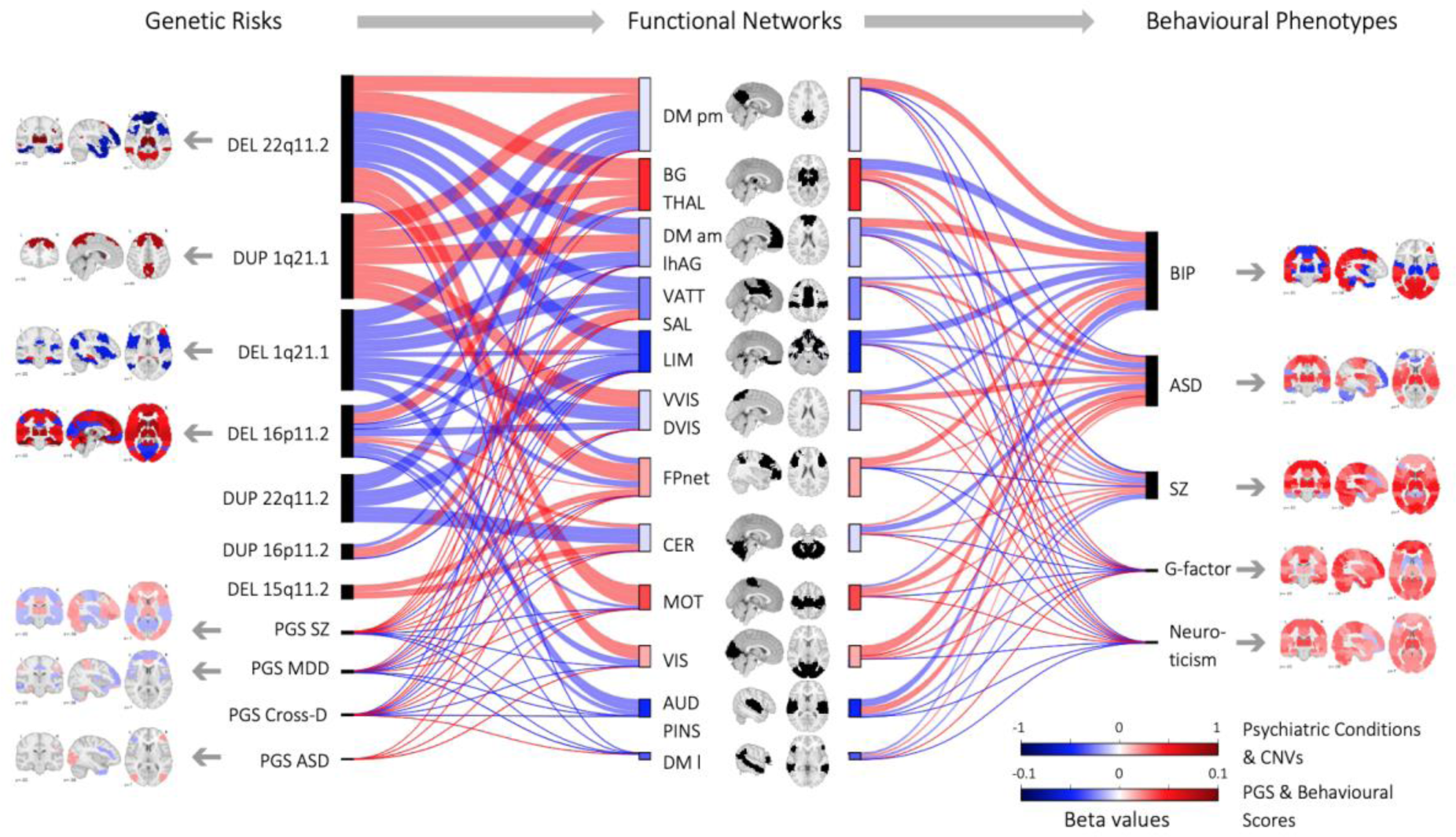
Effect size of dysconnectivity across 12 functional networks for CNVs, PGS, psychiatric conditions, and traits. Legend: Brain maps represent max beta value per seed-region for each FC-profile with connections surviving FDR (Table 2). Red = overconnectivity ; blue underconnectivity. color scale represents the beta value (z-score). The Sankey plot (middle of the Figure) shows dysconnectivity across 12 networks for genetic risk (left), conditions, and traits (right). The thickness of the connecting lines represents effect sizes (mean beta value of all FDR connections between a network and the rest of the brain). The length of rectangles represents the sum of effect sizes, which penalizes groups with higher statistical power. Abbreviations: DEL=Deletion, DUP=Duplication, ASD: autism spectrum disorder; SZ: schizophrenia; BIP: Bipolar disorder; PGS: Polygenic score; MDD: major depressive disorder; CrossD: Cross-Disorder.

### A landscape of functional correlation across genetic risk, psychiatric conditions, and traits

We further investigated the correlations between FC profiles presented above. This analysis was limited to the 20 whole-brain FC-profiles with significantly altered connections (FDR, **Table 2**). 84 out of 190 pairs of FC profiles showed correlations above what is expected by chance and 48 survived FDR (10,000 null correlations, **Figure 5AE**). Although, for the most part, genetic risk, diseases, and traits presented singular FC profiles (rFC < 0.5), the correlation matrix revealed a primary cluster of shared connectivity between ASD, SZ, BIP, 16p11.2, 1q21.1, and 22q11.1 deletions and duplications, 15q11.2 deletion, as well as neuroticism, PGS-MDD, PGS-ASD, and PGS-SZ. This “neuropsychiatric cluster” was negatively correlated with cluster 2 (‘cognition cluster’) driven by G-factor, Fluid intelligence, PGS-IQ, CT, SA, and PGS-SA.

### Thalamo-sensorimotor alterations are shared across CNVs, PGS, and idiopathic conditions

To characterize networks underlying clusters, we performed a principal component analysis (PCA) across the 14 FC-profiles encompassed in the neuropsychiatric cluster.

Two dimensions explained respectively 26% and 10% of the variance of the FC profiles. Dimension 1 was dominated by increased connectivity involving the somatomotor and thalamus-basal ganglia networks (**Figure 5B**). Dimension 2 was characterized by decreased connectivity between the visual network, the posterior-medial DMN, and the ventral attention and salience networks (**Figure 5C**). Because neuroticism and psychiatric conditions showed higher loadings on dimension 1 than CNVs (**Figure 5D**), we also performed a second PCA on CNVs separately, demonstrating that similar networks and connections were contributing to the main dimension (*r*=0.73).

**Figure 5.**
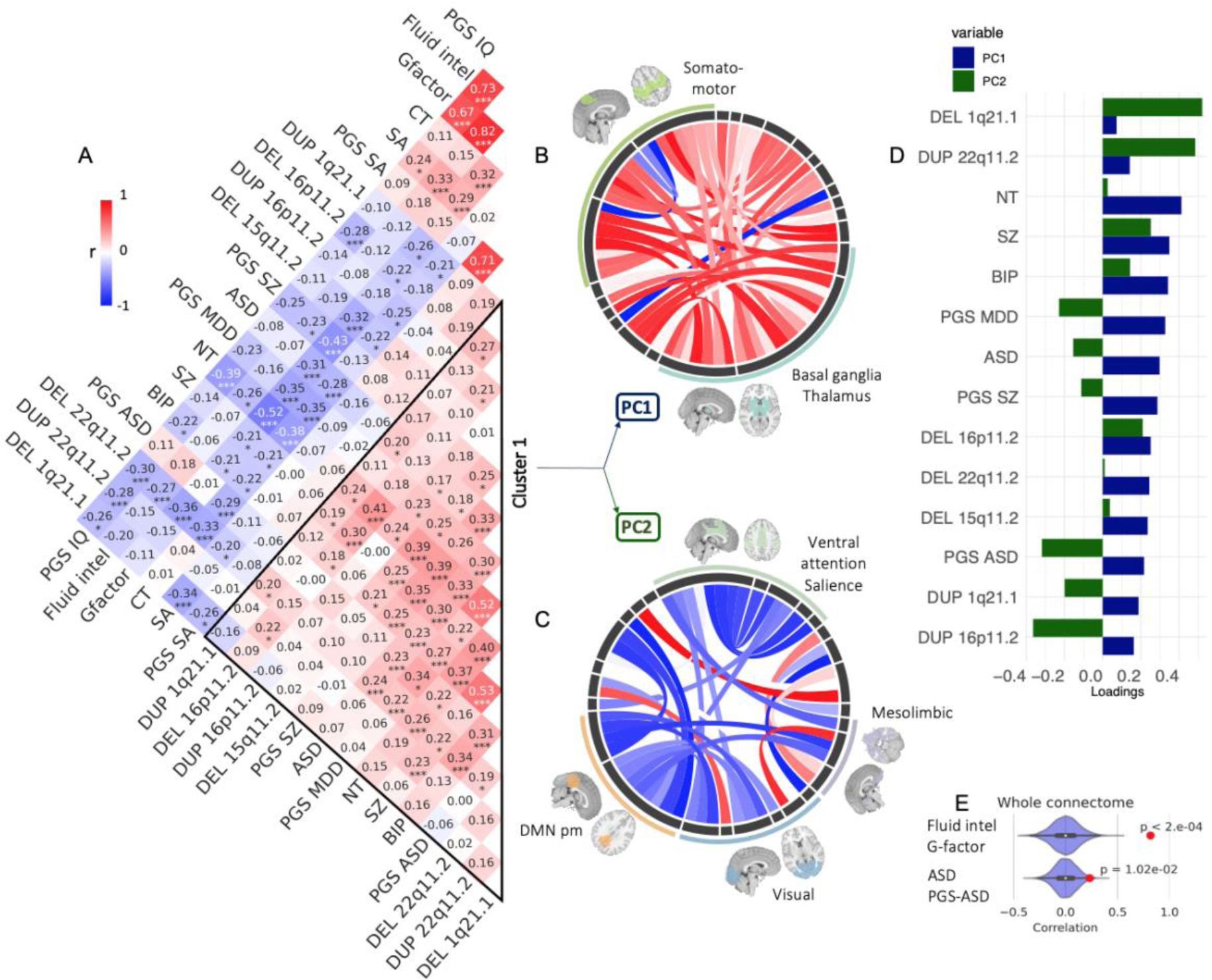
Atlas of functional connectivity relationships across psychiatric conditions, genetic risks and traits. Legend: A. Pearson correlation between 20 FC-profiles (2080 beta values from 20 CWAS). Stars represent significant correlations (* p<0.05, ** p<0.005, *** q FDR). Cluster 1 was defined by hierarchical clustering. B-D. PCA conducted on cluster 1. B-C Loadings of functional connections on PC1 (C) and PC2 (D) (overconnectivity in red, underconnectivity in blue). Each chord diagram shows the top 10% of connections. All 64 seed regions are represented in the black inner circle. Seed regions are grouped into functional networks. The width of the seed region in the black inner circle corresponds to the contribution of regions to the PC. Dimension 1 was dominated by overconnectivity of the thalamus, basal ganglia, and the somatomotor network. Dimension 2 was dominated by decreased connectivity between the visual network, the posterior-medial DMN, and the ventral attention and salience networks. D. Loadings of conditions and traits from cluster 1 on PC1 (blue) and PC2 (green) explaining respectively 26 and 10% of the connectome-wide variance across cluster 1. E. Density plots show examples of null distributions of correlations used to determine significance. FC profiles of Fluid Intelligence and G factor have the highest correlation overall, and those of ASD and PGS ASD have the lowest correlation that survives FDR. Abbreviations: ASD: autism spectrum disorder, SZ: schizophrenia, BIP: bipolar disorder, MDD: major depressive disorder, NT: Neuroticism, PGS: Polygenic score, Del: deletion, Dup: duplication, Fluid intel: fluid intelligence, SA: surface area, CT: cortical thickness, IQ: intelligence quotient; DMN pm: Default mode network posteromedial.

The regional FC profiles of the thalamus and dorsolateral motor network showed, as expected, similar clusters with much higher similarities among genetic risk, conditions, and traits (24 and 60 out of 190 correlations survived FDR respectively) (**Figure 6A** and Supplemental Figure 4).

**Figure 6.**
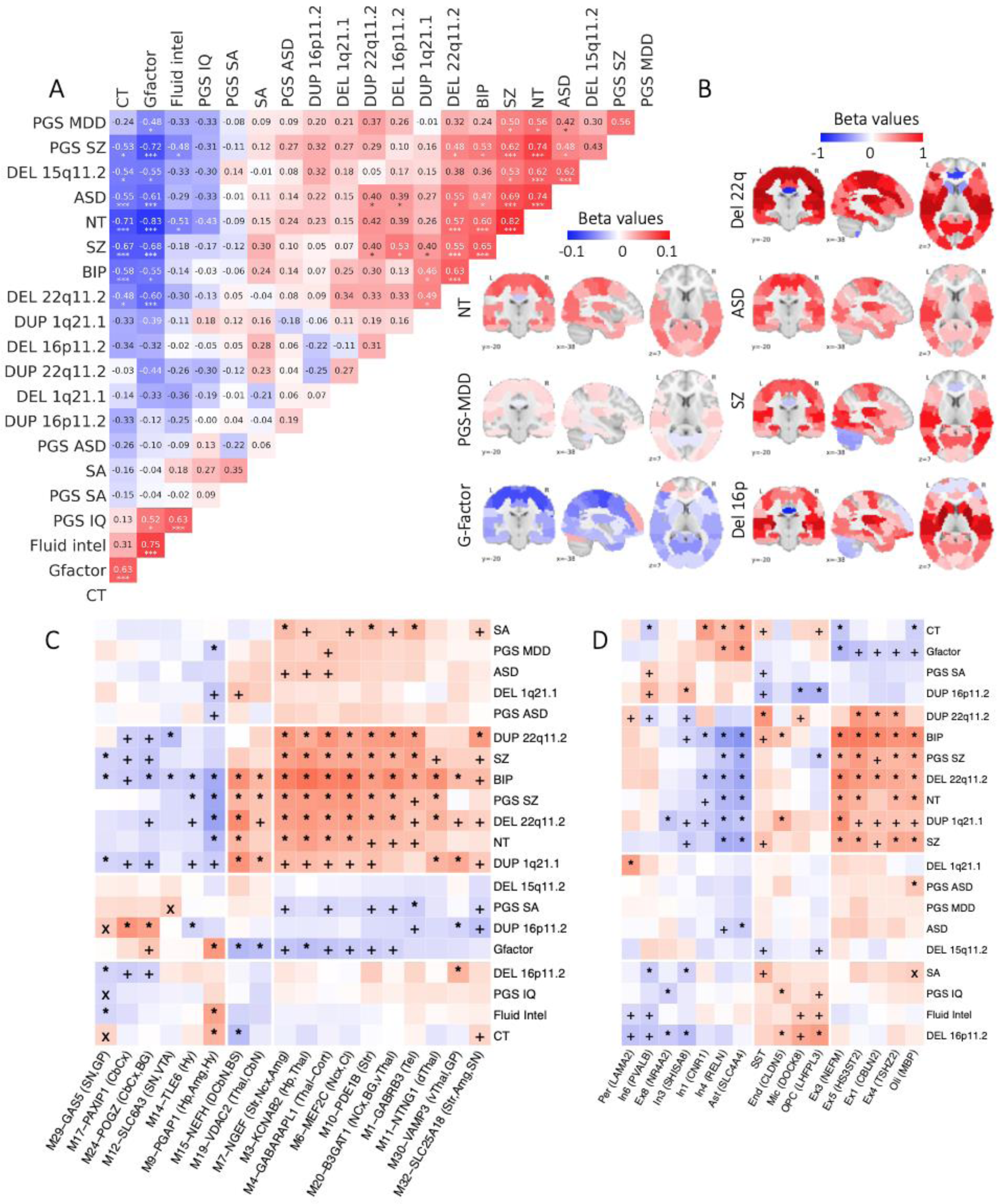
Relationship between thalamic dysconnectivity, transcription modules and brain cell types. Legend: A. Pearson correlation between 20 FC-profiles of the thalamus (64 beta values from 20 CWAS). Stars represent significant correlations (* p<0.05, ** p<0.005, *** q FDR). 49 out of 190 pairs of FC profiles showed correlations above what is expected by chance and 24 survived FDR (10,000 null correlations). B. Brain maps represent thalamic FC profiles (64 beta values for each connection between the thalamus and all other functional regions). Red = overconnectivity; blue underconnectivity. color scale represents the beta value (z-score). C-D: Heatmaps showing spatial relationship (Pearson correlation) between 20 FC-profiles and patterns of gene expression for C) AHBA co-expression module eigengenes (AHBA Nat Neuro 2015), D) CellType marker genes (29, 31); and CellType categories with marker gene name and AHBA modules with eigengene and anatomical associations are included in column names. Rows and columns are clustered using k-means. Significance (nominal p-values < 0.05) for the correlation, obtained using 10000 BrainSMASH surrogate profiles, are shown with “x”; LabelShuffle are shown with “+”; and Both with “*”. CellTypes categories include five interneuron subtypes (i.e., Somatostatin (SST), In1, In3, In4, and In6), five excitatory neuron subtypes (i.e., Ex1, Ex3, Ex4, Ex5, and Ex8), and six non-neuronal subtypes (Ast: Astrocytes; End: Endothelial; Mic: Microglia; Oli: Oligodendrocytes; OPC: oligodendrocyte precursor cells; and Per: Pericytes). Ex: Excitatory; In: Inhibitory. Abbreviations: ASD: autism spectrum disorder, SZ: schizophrenia, BIP: bipolar disorder, MDD: major depressive disorder, NT: Neuroticism, PGS: Polygenic score, Del: deletion, Dup: duplication, Fluid intel: fluid intelligence, SA: surface area, CT: cortical thickness. ABHA Module Anatomy abbreviations: Amg: Amygdala; BG: Basal Ganglia; BS: BrainStem; CbCx: Cerebellar Cortex; CbN: Cerebellar Nuclei; Cl: Claustrum; DCbN: Deep Cerebellar Nuclei; dThal: Dorsal Thalamus; Hp: Hippocampus; Hy: Hypothalamus; Ncx: Neocortex; SN: Substantia Nigra; Str: Striatum; Tel: Telencephalon; Thal: Thalamus; Thal-Cort: Thalamocortical; vThal: Ventral Thalamus; VTA: Ventral Tegmental Area. DEL: deletion; DUP: duplication.

### Thalamic overconnectivity is spatially associated with glutamatergic thalamic neurons

Finally, we sought to identify transcriptomic or cellular correlates of the thalamic over-connectivity observed across genetic risk and conditions. Towards this goal, we took advantage of gene co-expression relationships defined in a brain-wide laser-capture microdissection-microarray gene expression analysis conducted by the Allen Institute for Brain Science (AHBA) (*17*). Eigengenes of co-expression modules were correlated with the 20 thalamic FC-profiles. The ‘neuropsychiatric’ cluster was significantly correlated with transcriptional modules involving the thalamus (ventral, dorsal, and thalamocortical), striatum, and neocortex, but negatively correlated with modules involving cerebellar cortex (M17, M24) (**Figure 6B**).

To further refine our analysis, we sought to predict which cell types could represent key drivers of positive correlation between AHBA transcriptomic modules and thalamic FC profiles. We took advantage of a multi-brain region single nucleus RNA sequencing dataset generated for the normal adult human cerebral cortex (*29*), which provides key cell type specific marker genes for glutamatergic (excitatory), GABAergic (inhibitory), and non-neuronal cells. Thalamic FC profiles of the neuropsychiatric cluster were significantly associated with marker genes of 4 glutamatergic cell types, oligodendrocytes, and somatostatin interneurons. In contrast, genes enriched in GABAergic neurons and astrocytes were negatively correlated with psychiatric FC-profiles. Together, these mapping efforts suggest that glutamatergic thalamic neurons strongly correlate with thalamic FC profiles shared across multiple conditions, and foreshadow future analyses incorporating brain-wide single-cell molecular signatures identified using cellular-resolution biochemical studies.

## Discussion

### Main findings

Using the largest connectome-wide characterization of rare and common genetic risk, idiopathic psychiatric conditions, and traits, we showed that:

1. Common and rare genetic risk factors for psychiatric conditions impacted most FC networks when sample sizes provided sufficient power. Effect-sizes of CNVs on FC were correlated and consistent with their effects on cognitive ability and risk for ASD-SZ;
2. Polygenicity had a profound impact on FC signals: As CNVs increased in size and number of deleted or duplicated genes, their effect size on FC did not increase in an additive fashion and rapidly tapered off. Accordingly, PGS have minute effects on FC compared to CNVs, including those with similar effect-sizes on IQ and risk for disease;
3. The level of overlap between conditions and traits was stable from genes to transcription to whole-brain FC.
4. Overlaps at the whole-brain FC level were mild to moderate across genetic risk, conditions, and traits. Functional alterations driving these similarities included overconnectivity of the thalamus and somatomotor networks. Thalamic FC profiles of conditions and genetic risk were spatially associated with marker genes of excitatory thalamic neurons and thalamic transcriptional modules.

### Even small levels of polygenicity increase heterogeneity at the functional connectivity level

Our results suggest that polygenicity has a profound impact on FC signals. Although CNVs encompassing more dosage-sensitive genes have a larger effect-size on FC, this effect appears non-linear and tapers off. The effect size normalized for the number of genes or dosage-sensitive genes declines rapidly for increasingly multigenic CNVs (by an order of magnitude).

In contrast, effect-sizes on IQ is a linear (additive) function of the number of dosage-sensitive genes encompassed in CNVs (i.e., a dosage-sensitive gene decreases IQ by about 3 points - regardless of the size and number of genes encompassed in deletions or duplications).

Multigenic CNVs may therefore represent heterogeneous combinations of relatively distinct FC profiles associated with each dosage-sensitive gene. This suggests that genes within a CNV or a polygenic score may cancel out each other’s effects on FC, leading to weaker effect-sizes. This is striking for PGS, which show much smaller effects on FC (roughly 6-fold) than those observed for CNVs even after matching for their effects on intelligence or risk for SZ and ASD. PGS will therefore require functional partitioning in order to observe the expected effects on brain traits. Polygenicity may therefore predominantly result in “poly- connectivity”, a scenario where thousands of ASD or SZ genomic risk variants lead to a diverse set of connectivity patterns associated with the conditions.

### Shared FC profiles across conditions and traits parallel genetic and transcriptomic overlap

Our results demonstrate a stable overlap between conditions and traits at the genomic, transcription, and large-scale connectivity levels. This suggests that a major component of FC-profile correlations reflects genetically-based biological processes (**Figure 1A**), consistent with the heritability of functional networks (*30, 31*). Similar concordance was previously reported for genetic and transcriptomic (*28, 32*), as well as genetic and cortical thickness, overlaps (*33*). Stable concordance from molecular to large-scale brain networks is less likely to indicate a strong mechanistic convergence at any level from micro- to macroscopic measures. In other words, distinct genomic variants may lead to largely distinct FC profiles.

### Genetic risks converge on the thalamus and somatomotor network

Although whole-brain FC profiles were largely specific with mild correlations (r<0.5), genetic risk, psychiatric conditions, and neuroticism formed an FC cluster driven by shared overconnectivity of the thalamus / basal ganglia and the somatomotor networks. The implication of the somatomotor and basal ganglia/thalamus network across genetic risk and psychiatric conditions is in line with previous transdiagnostic and single condition neuroimaging studies (*34, 35*).

These functional hubs may be highly sensitive to a broad range of genetic risks for neuropsychiatric conditions. This may be related to the fact that functional and structural measures of the thalamus, basal ganglia (*36*), and unimodal regions (ie somatomotor) show less interindividual variability and higher heritability compared to heteromodal regions (*31*). G-factor and fluid intelligence showed the opposite thalamic pattern. This is in line with prior functional MRI studies demonstrating that thalamocortical pathways are engaged in memory, attention, and mental representations (*37, 38*).

Thalamic FC profiles were spatially correlated with transcription modules implicating the thalamus, and genes highly expressed in excitatory neurons. The thalamus is composed of diverse inhibitory and excitatory neuronal types distributed across anatomically defined nuclei. Thalamocortical projection neurons are largely excitatory (*39*), and therefore our spatial correlation is consistent with a hypothesis that changes in the activity of these cells are the primary driver of the thalamic FC alterations identified in our study. We anticipate that future studies incorporating single-cell transcriptomic profiles from the thalamus and other subcortical structures in the human brain will further refine these hypotheses.

## Limitations

Small sample sizes did not allow us to accurately characterize connectivity alterations for several CNVs. However, when samples were sufficiently powered, FC profiles appear to be robust - as shown by their correlation with previously published results.

This multisite study including clinically and non-clinically ascertained cohorts may have introduced biases. Confounding factors include sex bias, age differences, and medication status, which may have influenced some of the results. However, carefully conducted sensitivity analyses, matching control groups for sex, site, age, motion, and excluding individuals with medications (in idiopathic psychiatric cohorts) provided similar results (Supplemental results).

Unknown confounders could introduce a correlation structure between FC profiles that matches SNP-level genetic correlations, but “non-brain” related conditions and traits used in our analyses as controls suggest that this is unlikely.

## Conclusion

This systematic rsfMRI investigation of genetic risks and conditions has important implications for the identification of large-scale brain mechanisms involved in schizophrenia and autism.

Currently, polygenic scores are too heterogeneous for meaningful neuroimaging studies. On the other hand, rare genomic disorders are associated with an extremely diverse landscape of brain profiles. Future studies will require both in-depth partitionings of polygenic scores as well as clustering of rare variants - based on relevant gene functions - to obtain mechanistically coherent subgroups of individuals.

Despite this complexity, molecular risks and psychiatric conditions effects on connectivity do highlight convergence on the thalamus and the somatomotor network. Such findings open optimistic avenues to delineate general mechanisms - amenable to intervention - across conditions and genetic risks.

## Materials and Methods

### Sample

We analyzed 32,988 individuals from nine datasets (Table 1, **Figure 1B**, Supplementary Materials and Methods).

CNVs carriers and controls

‘Genetics-first’ cohorts were recruited based on the presence of a CNV, regardless of symptomatology, four consortia (two out of four have never been published before): the Simons Variation in Individuals Project (VIP) consortium data (16p11.2 and 1q21.1 CNVs carriers) (*40*), the University of California, Los Angeles (22q11.2 CNVs carriers), the Montreal rare genomic disorder family project (MRG, CHU Sainte-Justine, Montreal, Canada), and the Define neuropsychiatric-CNVs Project (Cardiff, UK) (see Supplementary Materials and Methods for individual dataset descriptions).

CNVs were also identified in an unselected population (UK Biobank) (see Supplementary Materials and Methods for the CNV calling procedure and final sample description).

The 9 non-psychiatric CNVs (light grey font) were defined as variants without any previous association with psychiatric conditions in large cases control studies (*24–26, 41*).

Idiopathic psychiatric conditions and respective controls

Individuals with idiopathic ASD, SZ, ADHD, BIP and their respective controls were sampled from 4 multicenter datasets (*42–45*) (Supplementary Materials and Methods).

Individuals with idiopathic ASD and their respective controls were sampled from the ABIDE1 multicenter dataset (*43*). Individuals with idiopathic SZ and their respective controls were obtained from aggregated fMRI data of 10 studies, and from the UCLA Consortium for Neuropsychiatric Phenomics (CNP) (*42*). Individuals diagnosed with ADHD (DSM-IV) and their respective controls were obtained from the ADHD-200 dataset (*44*) and the CNP. Individuals with idiopathic BIP and their respective controls were also sampled from the CNP dataset (Supplementary Materials and Methods).

Each cohort analyzed in this study was approved by the research ethics review boards of the respective institutions. Signed informed consent was obtained from all participants or their legal guardians before participation. Secondary analyses of the listed datasets for the purpose of this project were approved by the research ethics review board at Sainte Justine Hospital. Imaging data were acquired with site-specific MRI sequences.

### Resting-state functional MRI Preprocessing and QC procedures

All datasets were preprocessed using the same parameters with the same Neuroimaging Analysis Kit (NIAK), an Octave-based open-source processing and analysis pipeline (*46*). Preprocessed data were visually controlled for quality of the co-registration, head motion, and related artefacts by three raters (Supplementary Materials and Methods).

### Computing connectomes

We segmented the brain into 64 functional seed-based regions defined by the multi-resolution MIST brain parcellation (*47*). FC was computed as the temporal pairwise Pearson’s correlation between the average time series of the 64 seed-based regions, and then Fisher-z transformed. The connectome of each individual encompassed 2,080 connectivity values: (63x64)/2 = 2016 region-to-region connectivity + 64 within seed-based region connectivity. We chose the 64 parcel atlas of the multi-resolution MIST parcellation as it falls within the range of network resolution previously identified to be maximally sensitive to FC alterations in neurodevelopmental disorders such as ASD(*48*). We corrected for multiple comparisons using a false discovery rate strategy (*49*).

Statistical analyses were performed in Python using the scikit-learn library (*50*). Analyses were visualized in Python and R. Code for all analyses and visualizations is available online through the GitHub platform with Jupyter notebook: https://github.com/claramoreau9/NeuropsychiatricCNVs_Connectivity

### Polygenic Scores via Bayesian regression and continuous shrinkage priors (PRS-CS)

PRS-CS (*51*) was used to infer posterior effects using only those SNPs for individuals of European ancestry (EUR) from the UKBB dataset that were also present in both the discovery GWAS summary statistics and an external 1000 Genomes EUR-ancestry linkage disequilibrium (LD) panel. Posterior effects were computed using discovery GWAS for MDD (*52*), SZ (*53*), ASD (*3*), total cortical SA,(*5*) average CT,(*5*) inflammatory bowel disease (IBD),(*54*) Low-Density Lipoprotein (LDL),(*55*) and Chronic Kidney Disease (CKD) (^*56*^) (Supplementary method). Note that the discovery GWAS used for IQ, MDD, and CKD included individuals from the UKBB. To ensure convergence of the underlying Gibbs sampler algorithm, we ran 25,000 Markov chain Monte Carlo iterations and designated the first 10,000 MCMC iterations as burn-in. The PRS-CS global shrinkage parameter was set to 0.01 when the discovery GWAS had a sample size that was less than 200,000; otherwise, it was learned from the data using a fully Bayesian approach. Default settings were used for all other PRS-CS parameters. The EUR posterior effects were fed into PLINK 1.9 (*57*) to produce raw PGS separately for the EUR and white British UKBB cohorts, and R (^*58*^) was used to standardize the PGS for each cohort to mean = 0 and SD = 1. Standardized PGS were then adjusted by regressing out the first ten within-ancestry PCs.

### Statistical analyses

#### Connectome-wide association studies (CWAS)

CWAS was conducted by linear regression at the connectome level, in which z-scored FC was the dependent variable and clinical status or continuous trait was the explanatory variable. Controls were included for all sites which included cases. FC was standardized (z-scored) based on the variance of the controls used for each CWAS. Models were adjusted for sex, scanning site, head motion, global signal (=‘GSA’), and age. We determined whether a connection was significantly altered by the clinical status effect by testing whether the β value (regression coefficient associated with the clinical status variable) was significantly different from 0 using a two-tailed *t*-test. This regression test was applied independently to each of the 2,080 functional connections. We corrected for the number of tests (2, 080) using the Benjamini-Hochberg correction for FDR at a threshold of *q* < 0.05 (*49*), following the previously published recommendations(*59*). Cohen’s *d* values can therefore be interpreted as z-scores.

Before GSA, we defined the effect size of global FC shift as the mean of the β values across all 2,080 connections.

After GSA, the mean of β values was equal to zero and overall FC effect-size of a genetic variant or condition was defined as the top decile of all 2080 β values.

We tested if the effect sizes were significantly different from zero by conducting a permutation test, shuffling the clinical status labels of the individuals included in each CWAS (using 5,000 permutations). We thus estimated a valid permutation-based *p*-value associated with the observed global FC shift and the observed FC effect size (*60*).

We performed 36 CWAS (**Figure 1B**):

1. comparing FC between cases and controls for 16 CNVs at the 15q11.2, 1q21.1, 2q13, 16p13.11, 13q12.12, 17p12, 16p11.2, 22q11.2, TAR-1q21.1, and 15q13.3 loci, as well as for 4 idiopathic psychiatric cohorts (ASD, SZ, BIP, and ADHD). FC was standardized (z-scored) based on the variance of the respective control group.
2. investigating the linear effect of 10 continuous polygenic scores: ASD, MDD, SZ, Cross-disorder, Surface Area, Cortical Thickness as well as three non-brain related control traits IBD, LDL, and CKD.
3. investigating the linear effect of 6 continuous traits provided by UK-Biobank: Freesurfer derivatives (SA, CT, and volume), Neuroticism, and cognitive scores (G-Factor, Fluid intelligence).

All continuous traits were normalized within the UKBB sample.

#### Concordance between functional, genetic, and transcriptomic correlation

We computed correlations of whole-brain connectome profiles across pairs of conditions and traits (Pearson correlation) using the 2,080 beta values of each CWAS.

We obtained genetic correlation (rG) values across pairs of conditions and traits (neuroticism (*61*), intelligence (*6*), cross-disorder (*2*), brain morphological traits (*5*)) from previously published GWAS and the database of genetic correlations across traits (ldsc.broadinstitute.org). We also obtained correlation values of transcriptomic profiles between 10 pairs of conditions (*28*).

We performed concordance analyses between correlation at the genetic (rG), and functional connectivity (rFC) levels as well as the transcriptomic (rT) and the FC (rFC) levels using DescTools R package (*58*). The bias correction factor quantifies how far the best fit line deviates from 45 degrees.

Atlas of functional connectivity correlations across genetic risk, traits, and conditions.

We computed Pearson correlations between the 20 out of 36 whole-brain FC-profiles with significantly altered connections (FDR-corrected). For the significance of correlations between FC profiles, we generated a null distribution of 10,000 correlation values for each pair of conditions and traits. These 10000 null correlations were computed using null FC profiles. The latter were obtained by conducting 5,000 CWAS after shuffling the clinical status or trait values.

To obtain a *p*-value, the correlation value was compared to the null distribution. We corrected for the number of correlations (n=190) using the Benjamini-Hochberg correction for FDR at a threshold of *q* < 0.05 (*49*).

#### Cluster extraction and Principal Component analysis

We defined 2 clusters using hierarchical clustering (*hclust* function from *stats* R package) on the 20 FC-profiles. To identify the FC networks driving the ‘neuropsychiatric’ cluster 1, we conducted a PCA on the 14 scaled FC-Profiles within cluster 1 using the *prcomp* function from *stats R* package. Functional connections with top decile loadings for principal components 1 and 2 (PC1, PC2) were represented on chord diagrams using the *circlize* R package (code available on Github).

#### Aligning gene expression maps and functional parcellation

To investigate the transcriptomic relationship of FC-profiles, we aligned gene expression values in the adult human brain from the AHBA dataset (*17*) to the MIST64 brain parcellation (*47*), as described in (*20, 47, 62, 63*).

#### AHBA co-expression modules and eigengenes

We tested the association between gene co-expression modules and FC-profiles by computing spatial correlation (Pearson r) between expression patterns of eigengenes of co-expression modules and FC-profiles. We used eigengenes for the 18 major AHBA co-expression modules (*17*). The significance of the spatial correlation was tested using label-shuffling (permutation test by performing similar contrasts in 5000 randomly sampled groups) and BrainSMASH (*64*). K-means clustering was used for grouping co-expression modules and FC-profiles (ComplexHeatmap package in R, consensus k-means clusters with 1000 iterations are reported).

#### Cell-Type classes and marker genes

We assessed the association between cell-type classes and FC-profiles by computing spatial correlation (Pearson *r*) between gene expression patterns of cell-type marker genes and FC-profiles. To be consistent with previous studies of rs-fMRI and AHBA gene expression data, we used marker genes for 16 transcriptionally defined cell classes previously used in MDD (*65*) and SZ investigations (*66*). The cell marker genes are defined based on differential expression in each cell type relative to others, using Cortical single-nucleus droplet-based sequencing (snDrop-seq) data from (*29*). The 16 cell classes include five interneuron subtypes (i.e., somatostatin (SST), In1, In3, In4, and In6), five excitatory neuron subtypes (i.e., Ex1, Ex3, Ex4, Ex5, and Ex8), and six non-neuronal subtypes (astrocytes, endothelial, microglia, oligodendrocytes, oligodendrocyte precursor cells, and pericytes). The significance of the spatial correlation was tested using label-shuffling and BrainSMASH. In addition, we clustered the cell-type classes and FC-profiles using k-means clustering from the ComplexHeatmap package in R (consensus k-means clusters with 1000 iterations are reported).

## Acknowledgments

### Author contributions

C.A.M., S.J., and P.B. designed the overall study and drafted the manuscript.

#### Analyses

C.A.M., S.U., and A.H. processed 90% of all the fMRI data and performed all imaging analyses.

K.K. performed all the gene expression analyses.

G.H., J-L. M., and E. D. performed the CNVs calling.

P.O. preprocessed the SZ data.

H.S. performed 3/4 of the UKBiobank fMRI preprocessing.

L.M.S. and L.A. computed the polygenic scores.

A.L. gave feedbacks on the statistics used in this manuscript

T.J.N. and D.S helped write sections on cell types and co-expression modules - and contributed to the interpretation of the gene expression results.

P.M.T., T.B., T.R., D.C.G., A.M, and C.E.B. contributed to the interpretation of the data and reviewed the manuscript.

#### Data collection

K.J, C-O.M, P.T., N.Y., E.D., S.L., recruited/scanned patients for the Montreal rare genomic disorder (MRG) family dataset.

C.E.B. provided the UCLA 22q.11.2 fMRI data.

D.E.J.L., M.J.O., M.B.M.B., J.H, and A.I.S. provided the Cardiff CNV fMRI data

All authors provided feedback on the manuscript.

### Data and materials availability

Data from UK Biobank was downloaded under the application 40980, and can be accessed via their standard data access procedure (see http://www.ukbiobank.ac.uk/register-apply). UK

Biobank CNVs were called using the pipeline developed in Jacquemont Lab, and described in https://github.com/labjacquemont/MIND-GENESPARALLELCNV. The final CNV calls are available from UK Biobank returned datasets (Return ID: 3104, https://biobank.ndph.ox.ac.uk/ukb/dset.cgi?id=3104).

ABIDE1, COBRE, ADHD200, CNP, 16p11.2 SVIP data are publicly available: http://fcon_1000.projects.nitrc.org/indi/abide/abide_I.html, http://schizconnect.org/queries/new, http://fcon_1000.projects.nitrc.org/indi/adhd200/, https://www.openfmri.org/dataset/ds000030/, https://www.sfari.org/funded-project/simons-variation-in-individuals-project-simons-vip/. The 22q11.2 UCLA raw data are currently available by request from the PI. Raw imaging data for the Montreal rare genomic disorder family dataset is going to be available on the LORIS platform in 2022. The Cardiff raw data is not publicly available yet, contact the PI for further information. Allen Human Brain Atlas is available from https://human.brain-map.org/. Single-cell expression data is available from the National Center for Biotechnology Information under the accession code GSE97942 (https://www.ncbi.nlm.nih.gov/geo/query/acc.cgi?acc=GSE97942).

All processed connectomes are available through a request to the corresponding authors. Code for all analyses and visualizations, beta values, and p-values for the 36 FC-profiles are available online through the GitHub platform with Jupyter notebook: https://github.com/claramoreau9/NeuropsychiatricCNVs_Connectivity

### Funding

This research was supported by Compute Canada (ID 3037 and gsf-624), the Brain Canada Multi investigator research initiative (MIRI), Canada First Research Excellence Fund, Institute of Data Valorization, Healthy Brain Healthy Lives (Dr. Jacquemont). Dr. Jacquemont is a recipient of a Canada Research Chair in neurodevelopmental disorders, and a chair from the Jeanne et Jean Louis Levesque Foundation. This work was supported by a grant from the Brain Canada Multi-Investigator initiative (Dr. Jacquemont) and a grant from The Canadian Institutes of Health Research (CIHR 400528, Dr. Jacquemont). The Cardiff CNV cohort was supported by the Wellcome Trust Strategic Award “DEFINE” and the National Centre for Mental Health with funds from Health and Care Research Wales (code 100202/Z/12/Z). The CHUV cohort was supported by the SNF (Maillard Anne, Project, PMPDP3 171331). Data from the UCLA cohort provided by Dr. Bearden (participants with 22q11.2 deletions or duplications and controls) was supported through grants from the NIH (U54EB020403), NIMH (R01MH085953, R01MH100900, 1U01MH119736, R21MH116473), and the Simons Foundation (SFARI Explorer Award). Finally, data from another study were obtained through the OpenFMRI project (http://openfmri.org) from the Consortium for Neuropsychiatric Phenomics (CNP), which was supported by NIH Roadmap for Medical Research grants UL1-DE019580, RL1MH083268, RL1MH083269, RL1DA024853, RL1MH083270, RL1LM009833, PL1MH083271, and PL1NS062410. Dr P. Bellec is a fellow (“Chercheur boursier Junior 2”) of the “Fonds de recherche du Québec - Santé”, Data preprocessing and analyses were supported in part by the Courtois foundation (Dr Bellec). This work was supported by Simons Foundation Grant Nos. SFARI219193 and SFARI274424. We thank all of the families at the participating Simons Variation in Individuals Project (VIP) sites, as well as the Simons VIP Consortium. We appreciate obtaining access to imaging and phenotypic data on SFARI Base. Approved researchers can obtain the Simons VIP population dataset described in this study by applying at https://base.sfari.org. We are grateful to all families who participated in the 16p11.2 European Consortium. Dr. P. Thompson was funded in part by the U.S. NIH grants R01MH116147, P41EB015922, R01MH111671, and U01 AG068057. Ms. Petra Tamer received the Canadian Institute of Health Research (CIHR) Scholarship.

## Supporting information

Supplementary Materials, Methods, and Results

## Data Availability

Data from UK Biobank was downloaded under the application 40980, and can be accessed via their standard data access procedure (see http://www.ukbiobank.ac.uk/register-apply). UK Biobank CNVs were called using the pipeline developed in Jacquemont Lab, and described in https://github.com/labjacquemont/MIND-GENESPARALLELCNV. The final CNV calls are available from UK Biobank returned datasets (Return ID: 3104, https://biobank.ndph.ox.ac.uk/ukb/dset.cgi?id=3104). ABIDE1, COBRE, ADHD200, CNP, 16p11.2 SVIP data are publicly available: http://fcon_1000.projects.nitrc.org/indi/abide/abide_I.html, http://schizconnect.org/queries/new, http://fcon_1000.projects.nitrc.org/indi/adhd200/, https://www.openfmri.org/dataset/ds000030/, https://www.sfari.org/funded-project/simons-variation-in-individuals-project-simons-vip/. The 22q11.2 UCLA raw data are currently available by request from the PI. Raw imaging data for the Montreal rare genomic disorder family dataset is going to be available on the LORIS platform in 2022. The Cardiff raw data is not publicly available yet, contact the PI for further information. Allen Human Brain Atlas is available from https://human.brain-map.org/. Single-cell expression data is available from the National Center for Biotechnology Information under the accession code GSE97942 (https://www.ncbi.nlm.nih.gov/geo/query/acc.cgi?acc=GSE97942). All processed connectomes are available through a request to the corresponding authors. Code for all analyses and visualizations, beta values, and p-values for the 36 FC-profiles are available online through the GitHub platform with Jupyter notebook: https://github.com/claramoreau9/NeuropsychiatricCNVs_Connectivity

