## Supplementary Materials, Methods, and Results for "Atlas of functional connectivity relationships across rare and common genetic variants, traits, and psychiatric conditions"

**Supplementary Materials and Methods** 1

Cohorts 1

SFARI SimonsVIP-dataset 1

UCLA 22q11.2-dataset 1

Montreal rare genomic disorder family dataset (MRG) 2

Cardiff DEFINE-dataset 2

Idiopathic ASD dataset - ABIDE1 3

Idiopathic schizophrenia dataset 3

Idiopathic ADHD dataset - ADHD200 4

UCLA Consortium for Neuropsychiatric Phenomics - CNP 4

UK-BioBANK dataset 4

CNVs calling procedure in UK-BioBANK 5

SNP Sample Size Determination for polygenic scores 5

Supplementary Figure 1. SNP Sample Size Determination 7

Supplementary Table 1. Discovery GWAS used to compute polygenic scores with PRS-CS 8

Resting-state fMRI Preprocessing 8

Quality Control - preprocessed rs-fMRI data 9

Supplementary Table 2. Psychiatric diagnoses and FSIQ 10

Supplementary Table 3. Psychiatric diagnoses in UK-Biobank 10

**Supplementary Results** 12

Supplementary Table 4. CWAS with and without global signal adjustment 12

CNVs sensitivity analyses 13

16p11.2 CNVs 13

22q11.2 CNVs 14

Supplementary Figure 2. Mean connectivity shifts in clinically and non-clinically ascertained 16p11.2 and 22q11.2 CNVs carriers 14

1q21.1 CNVs 15

Supplementary Figure 3. Mean connectivity shifts in clinically and non-clinically ascertained 1q21.1 CNVs carriers 15

Sensitivity analyses excluding female participants in idiopathic psychiatric conditions and controls 16

Sensitivity analyses excluding ASD and ADHD participants with medication 16

Supplementary Table 5. Genetic, transcriptomic and functional correlation values 17

Supplementary Figure 4. Dorsolateral Motor network 18

### Supplementary Materials and Methods

#### Cohorts

##### SFARI *SimonsVIP*-dataset

Copy number variants (CNV) carriers were clinically ascertained. Imaging data of 16p11.2 CNV carriers and typically developing controls were acquired by the Simons variation in individuals project (VIP) consortium *(1)* across 2 sites. We excluded 50 individuals from the analysis due to the insufficient quality of the imaging data. The final 16p11.2 sample includes 146 individuals: 16p11.2 deletion (n=25) and duplication (n=26) carriers, 1q21.1 deletion (n=7) and duplication (n=4) carriers, and extrafamilial controls (n=84). Over 90% of the deletion carriers and 69% of the duplication carriers met the criteria for at least one clinical psychiatric diagnosis. Control subjects were recruited from the general population (extra-familial subjects) and had no major DSM-V diagnosis. The 16p11.2 duplication group includes 2 individuals with a triplication.

##### UCLA *22q11.2*-dataset

CNV carriers were clinically ascertained. Imaging data of 22q11.2 CNV carriers and typically developing (TD) controls were acquired at the University of California, Los Angeles (UCLA). Patients were ascertained from the UCLA or Children's Hospital, Los Angeles Pediatric Genetics, Allergy/Immunology and/or Craniofacial Clinics. Demographically comparable TD comparison subjects were recruited from the same communities as patients via web-based advertisements and by posting flyers and brochures at local schools, pediatric clinics, and other community sites. Exclusion criteria for all study participants included significant neurological or medical conditions (unrelated to 22q11.2 mutation) that might affect brain structure, history of head injury with loss of consciousness, insufficient fluency in English, and/or substance or alcohol abuse or dependence within the past 6 months. The UCLA Institutional Review Board approved all study procedures and informed consent documents. Scanning was conducted on an identical 3 T Siemens Trio MRI scanner with a 12-channel head coil at the University of California at Los Angeles Brain Mapping Center or at the Center for Cognitive Neuroscience *(2)*. We excluded 25 individuals from the analysis due to the insufficient quality of the imaging data. The final 22q11.2 sample includes 22q11.2 BPA-BPD deletion (n=43) and BPA-BPD duplication (n=10) carriers and extrafamilial controls (n=43).

#### Montreal rare genomic disorder family dataset (MRG)

Cognitive and behavioral measures were collected in families with at least one child who carries a CNVs classified as pathogenic or VUS (variation of uncertain significance). CNV carriers and their first-degree relatives were ascertained neurodevelopmental disorder clinic at the Sainte Justine Montreal hospital, Quebec-Canada (MP-21-2016-946, 4165). The characteristics of this cohort reflect the criteria for Chromosomal microarray testing in the neurodevelopmental disorder clinic which include: intellectual disabilities, learning disabilities, autism spectrum disorder (ASD) as well as children with several comorbidities including attention deficit hyperactivity disorder (ADHD), speech and language disorders and developmental coordination disorders. The same assessments performed in first-degree relatives (carriers and non-carriers) allow adjusting for the effect of the additional genetic and environmental background. All the MRI scans have been performed at the Montreal Neurological Institute using the same scanner (Prisma 3T). The acquisition time for the resting-state sequence was 7 minutes. The neuroimaging protocol was designed by John D. Lewis and is available online: http://www.bic.mni.mcgill.ca/users/jlewis/BrainCanada/MCIN/

Data from this cohort include 16p11.2 deletion (n=7) and duplication (n=3) carriers, 1q21.1 deletion (n=5) and duplication (n=1) carriers, 15q11.2 (n=1) duplication carrier, 22q11.2 (n=1) duplication carrier, and intrafamilial controls (n=47) after exclusion of 16 individuals due to quality control criteria.

#### Cardiff *DEFINE*-dataset

The Cardiff CNV cohort was supported by the Wellcome Trust Strategic Award "DEFINE" and the National Centre for Mental Health with funds from Health and Care Research Wales. CNV carriers were clinically ascertained. MRI data were acquired on a 3 Tesla General Electric HDx MRI system (GE Medical Systems, Milwaukee, WI) using an eight-channel receive-only head RF coil (as described in *(3)*). The acquisition time for functional resting-state data was 7 minutes. Following parameters have been used: Repetition Time = 2000 ms, Echo time = 35 ms, Slice Thickness = 3.4 mm (eyes open, fixation cross). The full protocol has been developed for the 100 brains project. 16p11.2 deletion (n=1) carrier, 1q21.1 deletion (n=3) and duplication (n=1) carriers, 15q11.2 (n=1) deletion carrier, 22q11.2 (n=1) duplication carrier, and extrafamilial controls (n=8) were included after the exclusion of 4 individuals due to quality control criteria.

#### Idiopathic ASD dataset - *ABIDE1*

The ABIDE dataset *(4)* is an aggregate sample of different studies including imaging and behavioral data for individuals with an ASD diagnosis and typically developing peers matched for age. Due to the small number of females in the ABIDE dataset, we excluded female individuals. To better account for biases in connectivity estimation due to differences in recording sites, subject age, and scanner motion, we created age and motion-matched subsamples for each recording site in ABIDE of individuals that passed our quality control criteria. We then excluded recording sites with fewer than 20 individuals (10 ASD, 10 controls). Our final ABIDE sample thus includes 459 male individuals, 225 individuals with ASD, and 234 healthy controls, from 10 recording sites.

#### Idiopathic schizophrenia dataset

We used fMRI data retrospectively aggregated from 10 distinct sites and studies. Brain imaging multi-state data were obtained through either the SchizConnect and OpenfMRI data sharing platforms (http://schizconnect.org *(5)*; https://openfmri.org *(6)*) or local scanning at the University of Montréal. All patients were diagnosed with schizophrenia (SZ) according to DSM-IV or DSM-V criteria, as a function of the time of the study. Sites samples were obtained after subjects were selected in order to ensure even proportions of SZ patients and controls within each site (from N = 9 to N = 42 per group) and to reduce between-group differences with regards to gender ratio (74% vs. 75% males in patients and controls, respectively), age distribution (34 vs. 32 years old on average) and motion levels (averaged frame displacement: 0.16 vs. 0.14 mm). Such matching of SZ and control subjects was achieved based on propensity scores. In total, we retained 242 SZ patients and 242 healthy controls in statistical analyses. Depending on the study, positive and negative symptoms were assessed with either the Positive and negative syndrome scale (PANSS,*(7)*) or the Scales for the assessment of positive/negative symptoms (SAPS/SANS,*(8)*). In order to allow for group analyses, SAPS/SANS scores were converted into PANSS scores using published regression-based equations*(9)*.

#### Idiopathic ADHD dataset - *ADHD200*

We used data provided by the ADHD-200 consortium and the neuro bureau ADHD-200 Preprocessed repository (8 cohorts http://fcon_1000.projects.nitrc.org/indi/adhd200/ *(10)*). Data from seven sites were retained after the exclusion of 447 individuals. We included in our study a total of 518 subjects, 187 patients diagnosed with ADHD and 331 healthy controls.

#### UCLA Consortium for Neuropsychiatric Phenomics - CNP

We downloaded T1-weighted Anatomical MPRAGE and resting-state fMRI BOLD data on the OpenfMRI platform (ds000030, https://www.openfmri.org/dataset/ds000030/) *(11)*. Subjects were scanned across 2 sites (UCLA) - with a similar 3T Siemens Trio machine. We excluded 13 individuals after visual quality control. Data included in our study (n=237) encompassed healthy individuals (n=113) and individuals diagnosed with SZ (n=41), BIP (n=44), or ADHD (n=39). These diagnoses followed the Structured Clinical Interview for DSM-IV.

#### UK-BioBANK dataset

The UK biobank genomics data *(12)* were available for 31,225 individuals with MRI. After standard quality control procedure, we were able to add to our sample: 2q13 deletion (n=183) and duplication (n=88) carriers, 16p11.2 deletion (n=4) and duplication (n=6) carriers, 1q21.1 deletion (n=10) and duplication (n=13) carriers, 1q21.1 TAR duplication carriers (n=29), 15q11.2 deletion (n=103), and duplication (n=136) carriers, 15q13.3 duplication (n=190) carriers and non-carriers (n=30,185) from UK-Biobank.

#### CNVs calling procedure in UK-BioBANK

The individual DNA (blood samples) were genotyped using the Affymetrix Axiom Array and UK BiLEVE Axiom array with common 733,256 markers mapped on the human genome version hg19. The UK Biobank provided normalized signal data of Log R Ratio and B Allele Frequency for each marker, which was formatted into standard input data suitable for the most common CNV calling software. We developed a CNV calling workflow with the ability to compute per individual CNV calling in parallel. The pipeline is compatible with Unix architectures and optimized for low-resource computers. We implemented CNV calling procedures for PennCNV*(13)* and QuantiSNP *(14)*. Our protocol was executed on Compute Canada servers. The pipeline is available through GitHub (https://github.com/labjacquemont/MIND-GENESPARALLELCNV) and a QuantiSNP docker image version were also made available *(15)*. We used default parameters for both algorithms to harmonize the CNV calling procedure (SNP coverage ≥ 3, likelihood score ≥ 15, and CNV length ≥1000 nt). Results were merged using CNVision script ($n_{cnv}$= 97,252) *(16)*. We annotated CNV within the 4 loci (1q21.1, 15q11.2, 16p11.2, 22q11.2). Each detected CNV was visually checked with SnipPeep (http://snippeep.sourceforge.net/). We pooled these data with previous observations reported by Kendall et al 2017 *(17)*.

#### SNP Sample Size Determination for polygenic scores

Polygenic prediction via Bayesian regression and continuous shrinkage priors (PRS-CS) *(18)* requires a single SNP sample size as an input. Given that most of our discovery GWAS were actually meta-GWAS that were comprised of individual studies that varied in terms of their sample size and the SNPs they included, the sample size often varied considerably between SNPs. To account for this reality, we examined the distribution of SNP sample sizes in R and excluded SNPs that had sample sizes that were less than half of the maximum SNP sample size. Of the remaining SNPs, the median SNP sample size was used as the PRS-CS sample size input.

As an example, consider the Freeze 2 EUR PTSD GWAS produced by the Psychiatric Genomics Consortium (PGC).*(19)* This meta-GWAS includes 9,766,174 SNPs with effective sample sizes that range from 17,559.4 to 70,237.5 (Figure S1-A). Given that PRS-CS uses only those discovery GWAS SNPs in common with both the EUR LD panel and the dataset, we started by retaining only the 1,116,862 SNPs that were present in the EUR LD panel. These SNPs also had effective sample sizes ranging from 17,559.4 to 70,237.5 (Figure S1-B). After we removed SNPs with effective sample sizes that were less than 35,000, the remaining 1,113,044 SNPs had effective sample sizes that ranged from 38,250.5 to 70,237.5 (Figure S1-C), with a median of 70,237.5. This median value was truncated to 70,237 and used as the SNP sample size when we ran PRS-CS. We made similar sample size determinations for the other discovery GWAS.

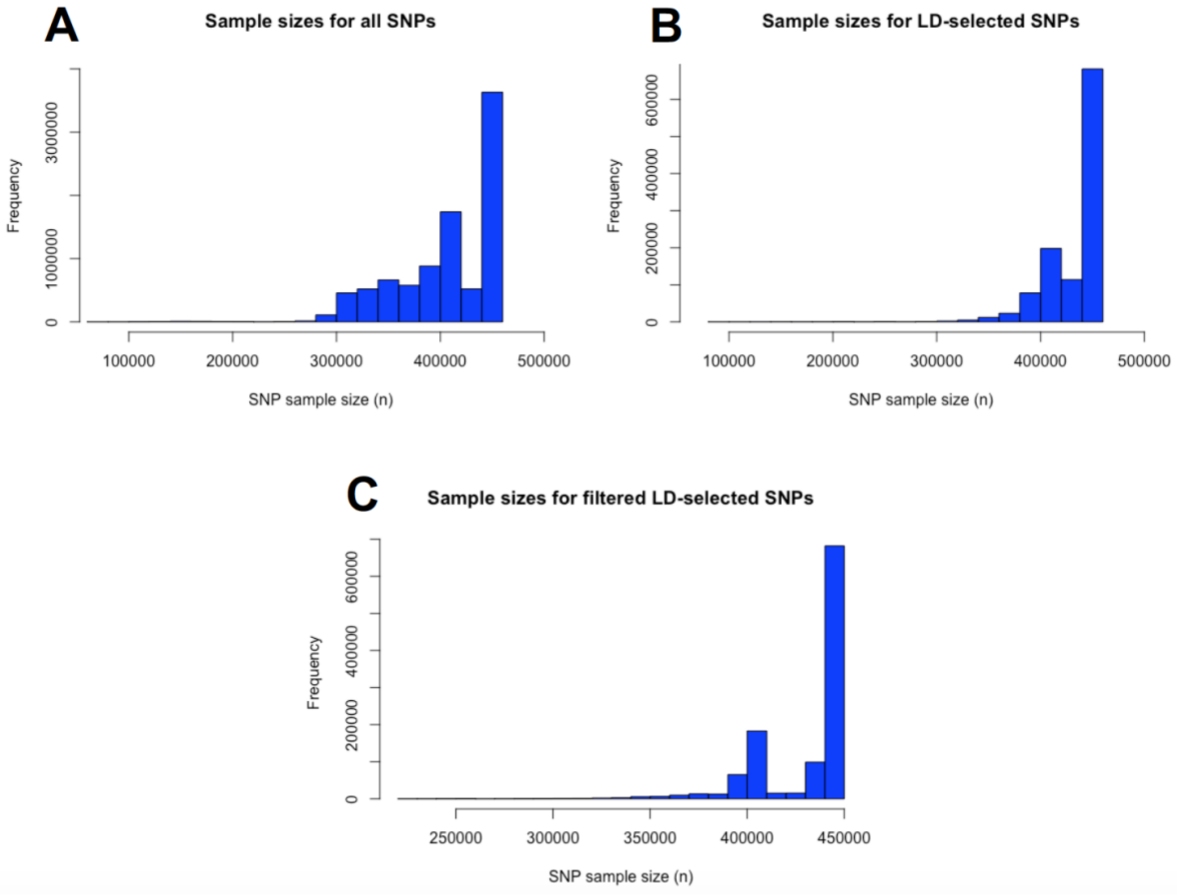

### Supplementary Figure 1. SNP Sample Size Determination

Legend: SNP sample size determination for the PGC PTSD Freeze 2 EUR meta-GWAS *(19)*. (A) This meta-GWAS contained 9,766,174 SNPs with effective sample sizes that ranged from 17,559.4 to 70,237.5. (B) The 1,116,862 SNPs in common with the PRS-CS EUR LD panel had this same range of effective sample sizes. (D) Filtering to retain only LD-selected SNPs with effective sample sizes of at least 35,000 resulted in 1,113,044 SNPs with effective sample sizes between 38,250.5 and 70,237.5. The median effective SNP sample size of 70,237.5 was truncated to 70,237 and used as the SNP sample size in PRS-CS.

### Supplementary Table 1. Discovery GWAS used to compute polygenic scores with PRS-CS

**Discovery GWAS used to compute polygenic scores with PRS-CS**

| Trait | Discovery GWAS | SNP Sample Size[a] (for PRS-CS) | SNP Count[b] (for PRS-CS) |
|---|---|---|---|
| **MDD** | Howard et al. (2019)*(20)* | 500,199 | 1,086,563 |
| **SZ** | Ruderfer et al. (2018)*(21)* | 87,491 | 1,079,332 |
| **ASD** | Grove et al. (2019)*(22)* | 46,350 | 1,094,054 |
| **SA** | Grasby et al. (2020)*(22)* | 32,176 | 1,101,464 |
| **CT** | Grasby et al. (2020)*(22)* | 32,872 | 1,102,328 |
| **IBD** | Liu et al. (2015)*(23)* | 34,762 | 1,105,053 |
| **LDL** | Willer et al. (2011)*(24)* | 89,856 | 977,929 |
| **CKD** | Wuttke et al. (2019)*(25)* | 438,997 | 1,111,519 |

MDD, major depressive disorder; SZ, schizophrenia; ASD, autism spectrum disorder; SA, total cortical surface area; CT, average cortical thickness; IBD, inflammatory bowel disease; LDL, low-density lipoprotein cholesterol; CKD, chronic kidney disease.

[a]PRS-CS requires a single SNP sample size; see Supplemental Figure 1.

[b]The "SNP count" is the number of SNPs in common between the discovery GWAS, the PRS-CS LD panel, and the genomic dataset; only these SNPs were used to calculate posterior effects.

### Resting-state fMRI Preprocessing

All datasets were preprocessed using the same parameters with the same Neuroimaging Analysis Kit (NIAK) version 0.12.4, an Octave-based open-source processing and analysis pipeline *(26)*. The first four volumes of each rs-fMRI time series were discarded to allow for magnetization to reach a steady state. Each data set was corrected for differences in slice acquisition time. Head motion parameters were estimated by spatially re-aligning individual timepoints with the median volume in the time series. This reference median volume was then aligned with the individual anatomical T1 image, which in turn was co-registered onto the MNI152 template space using an initial affine transformation, followed by a nonlinear transformation. Finally, each individual time point was mapped to the MNI space *(27)* using the combined spatial transformations. Slow frequency drifts were modeled on the entire time series as discrete cosine basis functions with a 0.01 Hz high-pass cut-off. Timepoints with excessive in-scanner motion (greater than 0.5 mm framewise displacement) were then censored from the time series by removing the affected timepoint as well as the preceding and following two-time points *(28)*. Nuisance covariates were regressed from the remaining time series: the previously estimated slow time drifts, the average signals in conservative masks of the white matter and lateral ventricles, and the first principal components (95% energy) of the estimated six rigid-body motion parameters and their squares. Data were then spatially smoothed with a 3D Gaussian kernel (FWHM = 6mm).

#### Quality Control - preprocessed rs-fMRI data

Preprocessed data were visually controlled for the quality of co-registration, head motion, and related artifacts by three raters. Not all datasets were examined by the same raters, yet all raters followed the same standardized quality-control procedure *(29)*. If there was a co-registration failure of either the functional image to individual T1 or individual T1 to MNI template registration, we attempted a manual fix by changing the parameters of the preprocessing pipeline. Individuals were excluded from the analysis if co-registration errors could not be fixed. Individuals were also excluded from the analysis if the average framewise displacement after motion censoring exceeded 0.5 mm or if fewer than 40-time frames remained.

### Supplementary Table 2. Psychiatric diagnoses and FSIQ

| CNV | Status | n clin | FSIQ | ASD | ADHD | SZ | BIP |
|---|---|---|---|---|---|---|---|
| **1q21.1** | DEL | 15 | 97 (17) | 2 | 2 | 0 | 0 |
| | DUP | 6 | 88 (29) | 2 | 2 | 0 | 0 |
| **22q11.2** | DEL | 43 | 77 (14) | 23 | 19 | 3 | 0 |
| | DUP | 12 | 96 (20) | 4 | 5 | 0 | 0 |
| **16p11.2** | DEL | 28 | 87 (15) | 7 | 7 | 0 | 0 |
| | DUP | 29 | 90 (21) | 3 | 3 | 0 | 0 |
| **Idiopathic Psychiatric Conditions** | **SZ** | 283 | - | - | - | 242 | - |
| | **BIP** | 44 | - | - | - | - | 44 |
| | **ASD** | 225 | 104 (17) | 225 | - | - | - |
| | **ADHD** | 226 | 107 (14) | - | 289 | - | - |

Legend: Diagnoses information and full-scale intelligence quotient (FSIQ, mean (standard deviation)) for the clinically ascertained CNV carriers, and psychiatric conditions. n clin: number of participants clinically ascertained; DEL: deletion; DUP: duplication; SZ: schizophrenia, ASD: Autism Spectrum Disorder; ADHD: Attention-Deficit/Hyperactivity-Disorder, BIP: Bipolar disorder.

### Supplementary Table 3. Psychiatric diagnoses in UK-Biobank

| CNV | Status | F17.1 Harmful use | F20-F29 SZ | F30.0 Hypomania | F32 Depressive episode | F41.9 Anxiety disorder | F42 OCD | F03 Dementia | F84.5 Asperger's syndrome | F90.0 ADHD | F99 Mental disorder unspecified |
|---|---|---|---|---|---|---|---|---|---|---|---|
| **15q11.2** | DEL | 2 | 0 | 0 | 2 | 0 | 0 | 0 | 0 | 0 | 0 |
| | DUP | 0 | 0 | 0 | 2 | 3 | 0 | 1 | 0 | 0 | 0 |
| **15q13.3** | DUP | 0 | 0 | 1 | 1 | 0 | 0 | 0 | 0 | 0 | 0 |
| **2q13** | DEL | 1 | 0 | 0 | 0 | 3 | 0 | 0 | 0 | 0 | 0 |
| | DUP | 1 | 0 | 0 | 0 | 1 | 0 | 0 | 0 | 0 | 0 |
| **Controls** | CON | 69 | 6 | 1 | 97 | 44 | 2 | 0 | 1 | 0 | 0 |
| **1q21.1** | DEL | 0 | 0 | 0 | 0 | 0 | 0 | 0 | 0 | 0 | 0 |
| | DUP | 0 | 0 | 0 | 1 | 0 | 0 | 0 | 0 | 1 | 1 |
| | DUP-TAR | 0 | 0 | 0 | 1 | 1 | 0 | 0 | 0 | 0 | 0 |
| **22q11.2** | DUP | 0 | 0 | 0 | 1 | 0 | 0 | 0 | 0 | 0 | 0 |
| **16p11.2** | DEL | 0 | 0 | 0 | 1 | 0 | 0 | 0 | 0 | 0 | 0 |
| | DUP | 0 | 0 | 0 | 0 | 0 | 0 | 0 | 0 | 0 | 0 |

Legend: Diagnosis information for the CNV carriers identified in the UK Biobank. More information about diagnostic codes is available on UK Biobank website.

### Supplementary Results

#### Supplementary Table 4. CWAS with and without global signal adjustment

| Metrics | | No global signal adjustment | | | | | | Global signal adjustment | | | | | |
|---|---|---|---|---|---|---|---|---|---|---|---|---|---|
| | | Connections | | Beta values | | mean | *pvalue* shift | Connections | | Beta values | | Mean top-dec | *pvalue* ES |
| | | *pos* | *neg* | *min* | *max* | | | *pos* | *neg* | *min* | *max* | | |
| CNV Deletion | 1q21.1 | 0 | 1 | -1.08 | 0.61 | -0.02 | ns | 1 | 11 | -1.07 | 0.62 | 0.44 | 0.002 |
| | 2q13 | 0 | 0 | -0.11 | 0.22 | 0.04 | ns | 0 | 0 | -0.15 | 0.19 | 0.11 | ns |
| | 13q12.12 | 0 | 0 | -0.52 | 0.53 | 0.03 | ns | 0 | 0 | -0.54 | 0.5 | 0.34 | ns |
| | 15q11.2 | 0 | 0 | -0.26 | 0.37 | 0.04 | ns | 1 | 0 | -0.29 | 0.36 | 0.2 | 0.01 |
| | 16p11.2 | 183 | 9 | -0.84 | 1.82 | 0.18 | 0.04 | 124 | 149 | -0.98 | 1.67 | 0.57 | <2e-4 |
| | 17p12 | 0 | 0 | -0.79 | 0.35 | -0.11 | ns | 0 | 0 | -0.67 | 0.46 | 0.40 | ns |
| | 22q11.2 | 0 | 38 | -1.6 | 0.80 | -0.25 | 0.04 | 4 | 13 | -1.48 | 1 | 0.65 | <2e-4 |
| CNV Duplication | TAR | 0 | 0 | -0.54 | 0.42 | -0.08 | ns | 0 | 0 | -0.48 | 0.51 | 0.28 | ns |
| | 1q21.1 | 102 | 0 | -0.39 | 1.12 | 0.30 | 0.008 | 4 | 0 | -0.62 | 0.84 | 0.48 | 0.002 |
| | 2q13 | 0 | 0 | -0.37 | 0.24 | -0.03 | ns | 0 | 0 | -0.34 | 0.26 | 0.18 | ns |
| | 13q12.12 | 0 | 0 | -0.31 | 0.79 | 0.31 | 0.009 | 0 | 0 | -0.53 | 0.48 | 0.31 | ns |
| | 15q11.2 | 0 | 0 | -0.28 | 0.18 | -0.06 | ns | 0 | 0 | -0.24 | 0.24 | 0.16 | 0.04 |
| | 15q13.3 | 0 | 0 | -0.2 | 0.18 | 0.007 | ns | 0 | 0 | -0.20 | 0.18 | 0.11 | ns |
| | 16p11.2 | 0 | 3 | -1.09 | 0.48 | -0.07 | ns | 4 | 3 | -1.04 | 0.55 | 0.38 | 0.002 |
| | 16p13.11 | 0 | 0 | -0.55 | 0.33 | -0.16 | 0.03 | 0 | 0 | -0.42 | 0.40 | 0.26 | ns |
| | 22q11.2 | 0 | 0 | -0.55 | 0.93 | 0.24 | 0.03 | 0 | 2 | -0.78 | 0.69 | 0.43 | 0.04 |
| Polygenic score | Cross Dis | 1 | 0 | -0.02 | 0.03 | -0.002 | ns | 23 | 22 | -.02 | .03 | 0.01 | <2e-4 |
| | ASD | 0 | 0 | -0.02 | 0.02 | 0 | ns | 3 | 1 | -.02 | .02 | 0.01 | ns |
| | Schizophrenia | 1 | 22 | -0.03 | 0.02 | -0.005 | ns | 30 | 27 | .02 | .03 | 0.01 | <2e-4 |
| | MDD | 0 | 2 | -0.02 | 0.02 | 0 | ns | 6 | 21 | -.02 | .02 | 0.01 | 0.003 |
| | IQ | 68 | 3 | -0.02 | 0.03 | 0.004 | ns | 74 | 42 | -.02 | .02 | 0.01 | <2e-4 |
| | LDL | 0 | 0 | -0.02 | 0.01 | -0.003 | ns | 0 | 0 | -.02 | .02 | 0.009 | ns |
| | IBD | 0 | 0 | -0.02 | 0.02 | 0 | ns | 1 | 1 | -.02 | 0.02 | 0.01 | ns |
| | CKD | 0 | 0 | -0.02 | 0.02 | 0 | ns | 0 | 0 | -.02 | .02 | 0.01 | ns |
| | Surface area | 0 | 0 | -0.02 | 0.03 | 0.002 | ns | 18 | 20 | -.03 | .03 | 0.02 | 0.0008 |
| | Thickness | 0 | 0 | -0.02 | 0.02 | 0.002 | ns | 0 | 0 | -.02 | .02 | 0.01 | ns |
| Psychiatric conditions | ASD | 2 | 71 | -0.44 | 0.34 | -0.09 | 0.01 | 21 | 11 | -0.35 | 0.42 | 0.21 | <2e-4 |
| | Schizophrenia | 21 | 1263 | -0.62 | 0.32 | -0.21 | <2e-4 | 208 | 242 | -0.41 | 0.5 | 0.30 | <2e-4 |
| | Bipolar | 0 | 16 | -0.77 | 0.48 | -0.16 | 0.08 | 22 | 12 | -0.67 | 0.63 | 0.42 | <2e-4 |
| | ADHD | 0 | 0 | -0.19 | 0.26 | 0.04 | 0.21 | 0 | 0 | -0.22 | 0.22 | 0.15 | <2e-4 |
| Morpho metry | Thickness | 927 | 178 | -0.05 | -0.07 | 0.01 | <2e-4 | 557 | 525 | -.06 | .06 | 0.04 | <2e-4 |
| | Surface area | 1459 | 150 | -0.08 | 0.2 | 0.03 | <2e-4 | 711 | 734 | -.11 | .13 | 0.07 | <2e-4 |
| | Volume | 1447 | 182 | -0.11 | 0.15 | 0.03 | <2e-4 | 704 | 716 | -.13 | -.12 | 0.07 | <2e-4 |
| Cognitive scores | Fluid Intel | 932 | 39 | -0.03 | 0.05 | 0.01 | 0.0004 | 315 | 286 | -.04 | .04 | 0.02 | <2e-4 |
| | G-Factor | 1021 | 29 | -0.05 | 0.06 | 0.02 | <2e-4 | 340 | 288 | -.06 | .05 | 0.03 | <2e-4 |
| | Neuroticism | 23 | 782 | -0.04 | 0.03 | -0.01 | 0.002 | 208 | 205 | -.03 | .04 | 0.02 | <2e-4 |

Legend: The number of significantly altered connections (FDR corrected) for each connectome-wide association study (n=36) before Global signal adjustment (GSA). DEL: deletion; DUP: duplication; ASD: autism spectrum disorder; SZ: schizophrenia; ADHD: attention deficit hyperactivity disorder. min-max: minimum-maximum of z-scored beta values; top decile: top decile of beta values; Connection pos: number of positive connections surviving FDR; Connection neg: number of negative connections surviving. Abbreviations: DEL: deletion; DUP: duplication; SZ: schizophrenia, ASD: Autism Spectrum Disorder; ADHD: Attention-Deficit / Hyperactivity-Disorder, BIP: Bipolar disorder, MDD: Major Depression Disorder, CrossD: Cross-disorder, LDL: Low-Density Lipoprotein, IBD: Inflammatory Bowel Disease, CKD: Chronic Kidney Disease; SA: Surface Area, CT: Cortical Thickness, IQ: intelligence quotient.

All the *beta*-values, *p*-values, and *q*-values are available on Github for the 36 FC-profiles:
https://github.com/claramoreau9/NeuropsychiatricCNVs_Connectivity/tree/master/results_tables
Interactive representations on brain maps *(30)* are also available, with and without GSA:
https://claramoreau9.github.io/Braimaps_Github.html

### CNVs sensitivity analyses

#### 16p11.2 CNVs

FC profiles obtained before GSA were correlated with our previous study (r=0.70, for 16p11.2 deletion, r=0.83, CI=[0.82; 0.84] for 16p11.2 duplication) *(31)*. We performed a sensitivity analysis only including clinically ascertained 16p11.2 carriers (Montreal Brain Canada, Define Cardiff, Lausanne, and SVIP cohorts, supplementary Figure 2). FC-profiles were strongly correlated to our previously published findings (r=0.79, CI=[0.77; 0.8] for 16p11.2 deletion, r=0.87, CI=[0.86; 0.88] for 16p11.2 duplication) *(31)*.

#### 22q11.2 CNVs

The GSA FC profile was correlated with the previously published FC profile without GSA *(31)* r=0.86 (CI=[0.85; 0.87]).

We performed a sensitivity analysis for 22q11.2 duplication, showing that FC-profiles using the full dataset and the FC-profiles excluding the non-clinically ascertained participants (UK-Biobank scanning sites). This new FC profile computed using UCLA, Brain Canada, and Define data, was highly correlated to previous findings (r=0.87, Supplementary Figure 2 *(31)*). We performed an additional sensitivity analysis matching controls for scanning site, age, sex, and motion (n=246) using the "matchControls" function in the {e1071} R package *(32)*. FC-profiles before and after matching were strongly correlated (r=0.96, CI=[0.95;0.97]).

### Supplementary Figure 2. Mean connectivity shifts in clinically and non-clinically ascertained 16p11.2 and 22q11.2 CNVs carriers

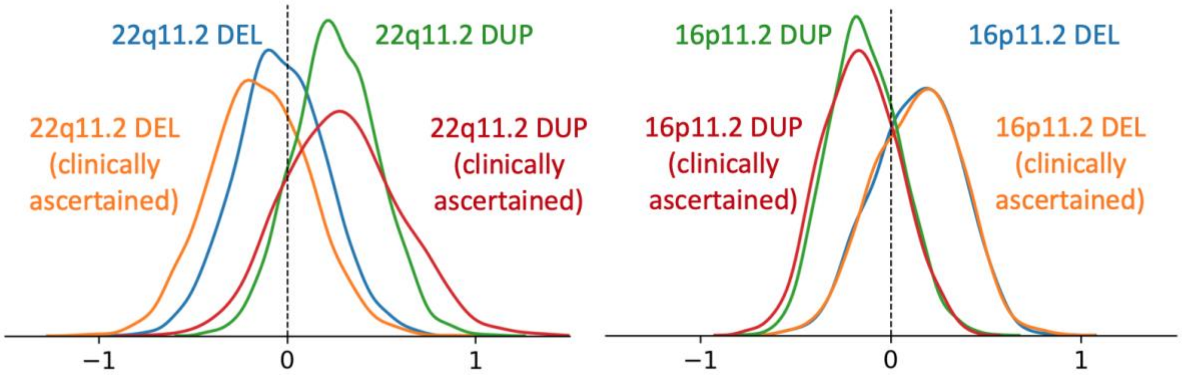


Legend: Density plots represent the FC profile distribution (2080 beta estimates from whole-brain contrast of cases versus controls) for the 22q11.2 CNVs (left) and 16p11.2 CNVs (right) groups, using all the available subjects (blue and green, for deletion and duplication respectively) or only the clinically ascertained CNVs carriers (orange and red).

##### 1q21.1 CNVs

Sensitivity analyses showed that FC-profiles obtained using all sites and only those including clinically ascertained carriers were correlated (r=0.75 for 1q21.1 deletion, and r=0.78 for 1q21.1 duplication, supplementary Figure 3).

#### Supplementary Figure 3. Mean connectivity shifts in clinically and non-clinically ascertained 1q21.1 CNVs carriers

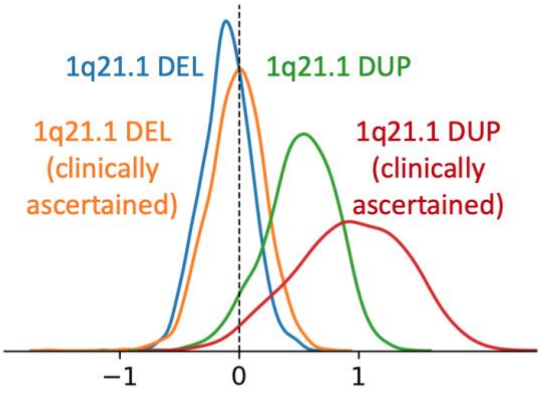


Legend. Density plots represent the distribution of 2080 beta estimates for the CWAS (whole brain contrast of cases versus controls) for 1q21.1 CNVs, using all the available subjects (blue and green, for deletion and duplication respectively) or only the clinically ascertained CNVs carriers (orange and red).

#### Sensitivity analyses excluding female participants in idiopathic psychiatric conditions and controls

A sensitivity analysis was performed on 210 male participants with SZ and 593 male controls. The beta map of this new CWAS analysis excluding females was highly correlated (r= 0.95) with the initial CWAS performed on the full sample. The same analysis was performed in the ADHD sample (n=160 male participants with ADHD, n=593 male controls). The beta map of the analysis excluding females was as well correlated (r= 0.80) with the initial CWAS performed on the full ADHD sample.

#### Sensitivity analyses excluding ASD and ADHD participants with medication

A sensitivity analysis was performed on 122 participants with ASD and the respective controls excluding subjects with medication at the MRI scan time and subjects without information about pharmacological treatment. The beta map of this new CWAS analysis was correlated (r= 0.81) with the initial CWAS performed on the full sample including subjects with medication. The same analysis was performed for ADHD using “Medication naive versus Not medication naive” information provided by the ADHD-200 consortium the same result (no connection survived FDR). We did not have individual medication information for the SZ cohorts.

### Supplementary Table 5. Genetic, transcriptomic and functional correlation values

| Pairs of conditions/traits | $r_G$ | $r_{FC}$ | $r_T$ | References $r_G$ | References $r_T$ |
|---|---|---|---|---|---|
| SZ BIP | 0.7 | 0.54 | 0.7 | Lee, P. H. et al. (2019) Table2.1 | Gandal, M. J. et al. (2018) TableS1.2 |
| SZ ASD | 0.22 | 0.3 | 0.476 | Lee, P. H. et al. (2019) Table2.1 | Gandal, M. J. et al. (2018) TableS1.2 |
| SZ ADHD | 0.13 | 0.12 | | Lee, P. H. et al. (2019) Table2.1 | |
| ASD BIP | 0.14 | 0.24 | 0.3389 | Lee, P. H. et al. (2019) Table2.1 | Gandal, M. J. et al. (2018) TableS1.2 |
| ASD ADHD | 0.37 | 0.03 | | Lee, P. H. et al. (2019) Table2.1 | |
| BIP ADHD | 0.14 | 0.15 | | Lee, P. H. et al. (2019) Table2.1 | |
| IQ ADHD | -0.27 | -0.08 | | Sniekers, S. et al. (2017) TableS13 | |
| IQ SZ | -0.2 | -0.06 | | Sniekers, S. et al. (2017) TableS13 | |
| IQ NT | -0.19 | -0.26 | | Sniekers, S. et al. (2017) TableS13 | |
| IQ BIP | -0.01 | -0.02 | | Sniekers, S. et al. (2017) TableS13 | |
| IQ ASD | 0.21 | -0.07 | | Sniekers, S. et al. (2017) TableS13 | |
| IQ ICV | 0.29 | 0.25 | | Sniekers, S. et al. (2017) TableS13 | |
| NT ADHD | 0.24 | 0.14 | | Nagel, M. et al. (2018) TableS37 | |
| NT SZ | 0.2 | 0.41 | | Nagel, M. et al. (2018) TableS37 | |
| NT ASD | 0.08 | 0.34 | | Nagel, M. et al. (2018) TableS37 | |
| NT BIP | 0.1 | 0.37 | | Nagel, M. et al. (2018) TableS37 | |
| NT ICV | -0.14 | 0.02 | | Nagel, M. et al. (2018) TableS37 | |
| SA NT | -0.129 | -0.07 | | Grasby, K. L. et al. (2020) TableS16 | |
| SA ADHD | -0.17 | 0.04 | | Grasby, K. L. et al. (2020) TableS16 | |
| SA IQ | 0.189 | 0.33 | | Grasby, K. L. et al. (2020) TableS16 | |
| SA BIP | 0.066 | 0 | | Grasby, K. L. et al. (2020) TableS16 | |
| SA SZ | -0.023 | 0 | | Grasby, K. L. et al. (2020) TableS16 | |
| SA ASD | 0.032 | -0.16 | | Grasby, K. L. et al. (2020) TableS16 | |
| SA ICV | 0.845 | 0.92 | | Grasby, K. L. et al. (2020) TableS9 | |
| CT ICV | -0.051 | 0.05 | | Grasby, K. L. et al. (2020) TableS9 | |
| CT ASD | -0.034 | -0.28 | | Grasby, K. L. et al. (2020) TableS16 | |
| CT ADHD | -0.024 | 0.1 | | Grasby, K. L. et al. (2020) TableS16 | |
| CT BIP | 0.04 | -0.24 | | Grasby, K. L. et al. (2020) TableS16 | |
| CT SZ | 0.008 | -0.21 | | Grasby, K. L. et al. (2020) TableS16 | |
| CT NT | 0.023 | -0.38 | | Grasby, K. L. et al. (2020) TableS16 | |
| CT IQ | 0.015 | 0.15 | | Grasby, K. L. et al. (2020) TableS16 | |
| PGS-MDD ASD | 0.45 | 0.29 | 0.0645 | Lee, P. H. et al. (2019) Table2.1 | Gandal, M. J. et al. (2018) TableS1.2 |
| PGS-MDD SZ | 0.34 | 0.22 | 0.237 | Lee, P. H. et al. (2019) Table2.1 | Gandal, M. J. et al. (2018) TableS1.2 |
| PGS-MDD BIP | 0.36 | 0.31 | 0.2423 | Lee, P. H. et al. (2019) Table2.1 | Gandal, M. J. et al. (2018) TableS1.2 |
| PGS-IBD PGSMDD | 0.02 | 0.08 | -0.036 | CDG of the PGC et al. (2013) TableS6 | Gandal, M. J. et al. (2018) TableS1.2 |
| PGS-IBD SZ | -0.01 | 0.01 | 0.008 | CDG of the PGC et al. (2013) TableS6 | Gandal, M. J. et al. (2018) TableS1.2 |
| PGS-IBD ASD | -0.07 | 0.05 | 0.21 | CDG of the PGC et al. (2013) TableS6 | Gandal, M. J. et al. (2018) TableS1.2 |
| PGS-IBD BIP | -0.05 | 0.08 | -0.06 | CDG of the PGC et al. (2013) TableS6 | Gandal, M. J. et al. (2018) TableS1.2 |

*Papers that computed $r_G$ and $r_T$ (22, 33–37) Abbreviations: ASD: autism spectrum disorder, SZ: schizophrenia, BIP: bipolar disorder, MDD: major depressive disorder, NT: Neuroticism, PGS: Polygenic score, Del: deletion, Dup: duplication, Fluid intel: fluid intelligence, SA: surface area, CT: cortical thickness, IQ: intelligence quotient.*

### Supplementary Figure 4. Dorsolateral Motor network

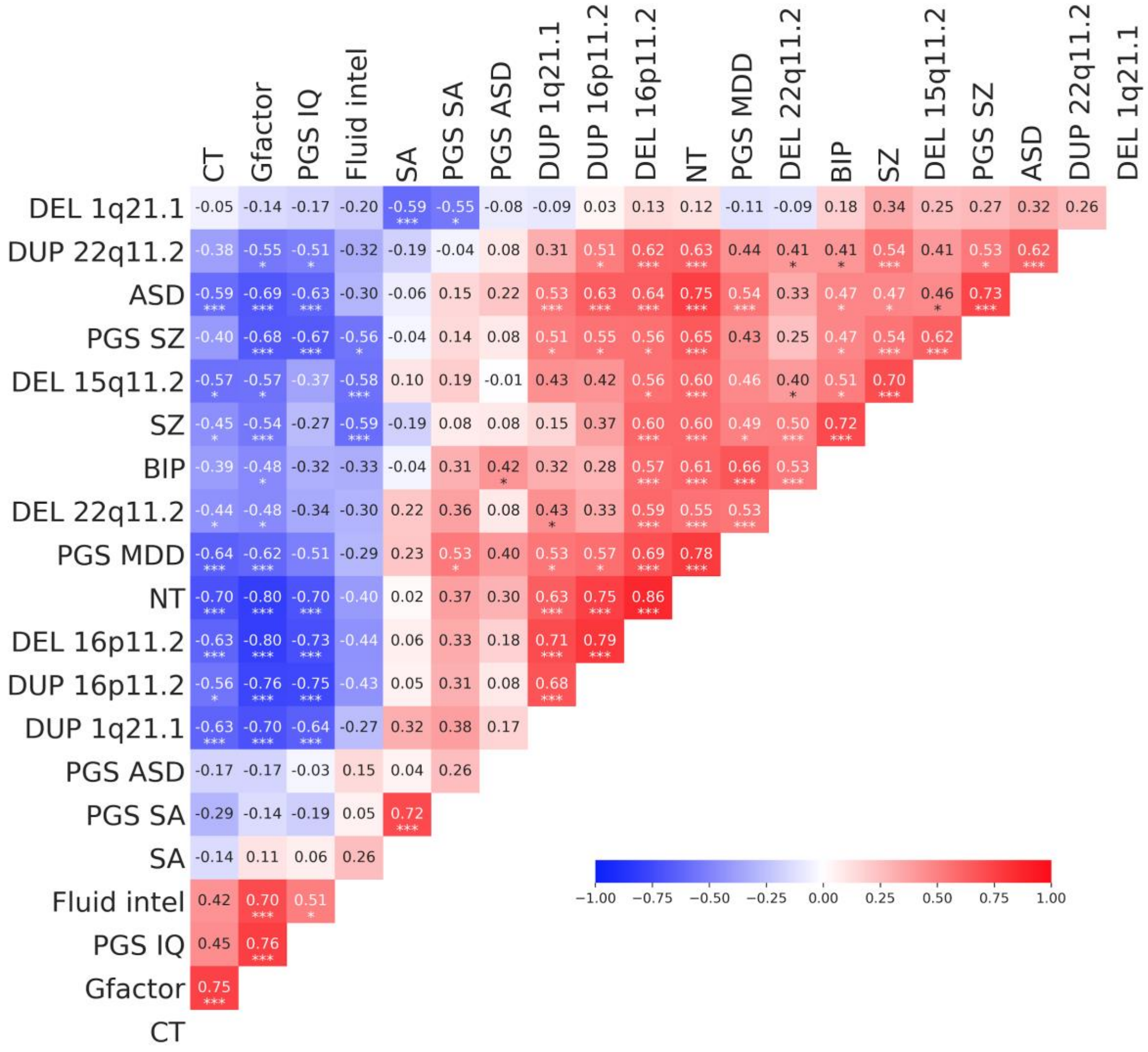


*Legend. Pearson correlation between 2 FC-profiles of the dorsolateral motor network (64 beta values from 20 CWAS). Stars represent significant correlations (* p<0.05, ** p<0.005, *** q FDR). 92 out of 190 pairs of FC profiles showed correlations above what is expected by chance and 60 survived FDR (10,000 null correlations). Abbreviations: ASD: autism spectrum disorder, SZ: schizophrenia, BIP: bipolar disorder, MDD: major depressive disorder, NT: Neuroticism, PGS: Polygenic score, Del: deletion, Dup: duplication, Fluid intel: fluid intelligence, SA: surface area, CT: cortical thickness, IQ: intelligence quotient*

19. C. M. Nievergelt, A. X. Maihofer, T. Klengel, E. G. Atkinson, C.-Y. Chen, K. W. Choi, J. R. I. Coleman, S. Dalvie, L. E. Duncan, J. Gelernter, D. F. Levey, M. W. Logue, R. Polimanti, A. C. Provost, A. Ratanatharathorn, M. B. Stein, K. Torres, A. E. Aiello, L. M. Almli, A. B. Amstadter, S. B. Andersen, O. A. Andreassen, P. A. Arbisi, A. E. Ashley-Koch, S. B. Austin, E. Avdibegovic, D. Babić, M. Bækvad-Hansen, D. G. Baker, J. C. Beckham, L. J. Bierut, J. I. Bisson, M. P. Boks, E. A. Bolger, A. D. Børglum, B. Bradley, M. Brashear, G. Breen, R. A. Bryant, A. C. Bustamante, J. Bybjerg-Grauholm, J. R. Calabrese, J. M. Caldas-de-Almeida, A. M. Dale, M. J. Daly, N. P. Daskalakis, J. Deckert, D. L. Delahanty, M. F. Dennis, S. G. Disner, K. Domschke, A. Dzubur-Kulenovic, C. R. Erbes, A. Evans, L. A. Farrer, N. C. Feeny, J. D. Flory, D. Forbes, C. E. Franz, S. Galea, M. E. Garrett, B. Gelaye, E. Geuze, C. Gillespie, A. G. Uka, S. D. Gordon, G. Guffanti, R. Hammamieh, S. Harnal, M. A. Hauser, A. C. Heath, S. M. J. Hemmings, D. M. Hougaard, M. Jakovljevic, M. Jett, E. O. Johnson, I. Jones, T. Jovanovic, X.-J. Qin, A. G. Junglen, K.-I. Karstoft, M. L. Kaufman, R. C. Kessler, A. Khan, N. A. Kimbrel, A. P. King, N. Koen, H. R. Kranzler, W. S. Kremen, B. R. Lawford, L. A. M. Lebois, C. E. Lewis, S. D. Linnstaedt, A. Lori, B. Lugonja, J. J. Luykx, M. J. Lyons, J. Maples-Keller, C. Marmar, A. R. Martin, N. G. Martin, D. Maurer, M. R. Mavissakalian, A. McFarlane, R. E. McGlinchey, K. A. McLaughlin, S. A. McLean, S. McLeay, D. Mehta, W. P. Milberg, M. W. Miller, R. A. Morey, C. P. Morris, O. Mors, P. B. Mortensen, B. M. Neale, E. C. Nelson, M. Nordentoft, S. B. Norman, M. O’Donnell, H. K. Orcutt, M. S. Panizzon, E. S. Peters, A. L. Peterson, M. Peverill, R. H. Pietrzak, M. A. Polusny, J. P. Rice, S. Ripke, V. B. Risbrough, A. L. Roberts, A. O. Rothbaum, B. O. Rothbaum, P. Roy-Byrne, K. Ruggiero, A. Rung, B. P. F. Rutten, N. L. Saccone, S. E. Sanchez, D. Schijven, S. Seedat, A. V. Seligowski, J. S. Seng, C. M. Sheerin, D. Silove, A. K. Smith, J. W. Smoller, S. R. Sponheim, D. J. Stein, J. S. Stevens, J. A. Sumner, M. H. Teicher, W. K. Thompson, E. Trapido, M. Uddin, R. J. Ursano, L. L. van den Heuvel,

M. Van Hooff, E. Vermetten, C. H. Vinkers, J. Voisey, Y. Wang, Z. Wang, T. Werge, M. A. Williams, D. E. Williamson, S. Winternitz, C. Wolf, E. J. Wolf, J. D. Wolff, R. Yehuda, R. M. Young, K. A. Young, H. Zhao, L. A. Zoellner, I. Liberzon, K. J. Ressler, M. Haas, K. C. Koenen, International meta-analysis of PTSD genome-wide association studies identifies sex- and ancestry-specific genetic risk loci, *Nat. Commun.* **10**, 4558 (2019).

20. D. M. Howard, M. J. Adams, T.-K. Clarke, J. D. Hafferty, J. Gibson, M. Shirali, J. R. I. Coleman, S. P. Hagenaars, J. Ward, E. M. Wigmore, C. Alloza, X. Shen, M. C. Barbu, E. Y. Xu, H. C. Whalley, R. E. Marioni, D. J. Porteous, G. Davies, I. J. Deary, G. Hemani, K. Berger, H. Teismann, R. Rawal, V. Arolt, B. T. Baune, U. Dannlowski, K. Domschke, C. Tian, D. A. Hinds, 23andMe Research Team, Major Depressive Disorder Working Group of the Psychiatric Genomics Consortium, M. Trzaskowski, E. M. Byrne, S. Ripke, D. J. Smith, P. F. Sullivan, N. R. Wray, G. Breen, C. M. Lewis, A. M. McIntosh, Genome-wide meta-analysis of depression identifies 102 independent variants and highlights the importance of the prefrontal brain regions, *Nat. Neurosci.* **22**, 343–352 (2019).

21. Bipolar Disorder and Schizophrenia Working Group of the Psychiatric Genomics Consortium. Electronic address:, Bipolar Disorder and Schizophrenia Working Group of the Psychiatric Genomics Consortium, Genomic Dissection of Bipolar Disorder and Schizophrenia, Including 28 Subphenotypes, *Cell* **173**, 1705–1715.e16 (2018).

22. K. L. Grasby, N. Jahanshad, J. N. Painter, L. Colodro-Conde, J. Bralten, D. P. Hibar, P. A. Lind, F. Pizzagalli, C. R. K. Ching, M. A. B. McMahon, N. Shatokhina, L. C. P. Zsembik, S. I. Thomopoulos, A. H. Zhu, L. T. Strike, I. Agartz, S. Alhusaini, M. A. A. Almeida, D. Alnæs, I. K. Amlien, M. Andersson, T. Ard, N. J. Armstrong, A. Ashley-Koch, J. R. Atkins, M. Bernard, R. M. Brouwer, E. E. L. Buimer, R. Bülow, C. Bürger, D. M. Cannon, M. Chakravarty, Q. Chen, J. W. Cheung, B. Couvy-Duchesne, A. M. Dale, S. Dalvie, T. K. de Araujo, G. I. de Zubicaray, S. M. C. de Zwarte, A. den Braber, N. T. Doan, K. Dohm, S. Ehrlich, H.-R. Engelbrecht, S. Erk, C. C. Fan, I. O. Fedko, S. F. Foley, J. M. Ford, M. Fukunaga, M. E. Garrett, T. Ge, S. Giddaluru, A. L. Goldman, M. J. Green, N. A. Groenewold, D. Grotegerd, T. P. Gurholt, B. A. Gutman, N. K. Hansell, M. A. Harris, M. B. Harrison, C. C. Haswell, M. Hauser, S. Herms, D. J. Heslenfeld, N. F. Ho, D. Hoehn, P. Hoffmann, L. Holleran, M. Hoogman, J.-J. Hottenga, M. Ikeda, D. Janowitz, I. E. Jansen, T. Jia, C. Jockwitz, R. Kanai, S. Karama, D. Kasperaviciute, T. Kaufmann, S. Kelly, M. Kikuchi, M. Klein, M. Knapp, A. R. Knodt, B. Krämer, M. Lam, T. M. Lancaster, P. H. Lee, T. A. Lett, L. B. Lewis, I. Lopes-Cendes, M. Luciano, F. Macciardi, A. F. Marquand, S. R. Mathias, T. R. Melzer, Y. Milaneschi, N. Mirza-Schreiber, J. C. V. Moreira, T. W. Mühleisen, B. Müller-Myhsok, P. Najt, S. Nakahara, K. Nho, L. M. Olde Loohuis, D. P. Orfanos, J. F. Pearson, T. L. Pitcher, B. Pütz, Y. Quidé, A. Ragothaman, F. M. Rashid, W. R. Reay, R. Redlich, C. S. Reinbold, J. Repple, G. Richard, B. C. Riedel, S. L. Risacher, C. S. Rocha, N. R. Mota, L. Salminen, A. Saremi, A. J. Saykin, F. Schlag, L. Schmaal, P. R. Schofield, R. Secolin, C. Y. Shapland, L. Shen, J. Shin, E. Shumskaya, I. E. Sønderby, E. Sprooten, K. E. Tansey, A. Teumer, A. Thalamuthu, D. Tordesillas-Gutiérrez, J. A. Turner, A. Uhlmann, C. L. Vallerga, D. van der Meer, M. M. J. van Donkelaar, L. van Eijk, T. G. M. van Erp, N. E. M. van Haren, D. van Rooij, M.-J. van Tol, J. H. Veldink, E. Verhoef, E. Walton, M. Wang, Y. Wang, J. M. Wardlaw, W. Wen, L. T. Westlye, C. D. Whelan, S. H. Witt, K. Wittfeld, C. Wolf, T. Wolfers, J. Q. Wu, C. L. Yasuda, D. Zaremba, Z. Zhang, M. P. Zwiers, E. Artiges, A. A. Assareh, R. Ayesa-Arriola, A. Belger, C. L. Brandt, G. G. Brown, S. Cichon, J. E. Curran, G. E. Davies, F. Degenhardt, M. F. Dennis, B. Dietsche, S. Djurovic, C. P. Doherty, R. Espiritu, D. Garijo, Y. Gil, P. A. Gowland, R. C. Green, A. N. Häusler, W. Heindel, B.-C. Ho, W. U. Hoffmann, F. Holsboer, G. Homuth, N. Hosten, C. R. Jack Jr, M. Jang, A. Jansen, N. A. Kimbrel, K. Kolskår, S. Koops, A. Krug, K. O. Lim, J. J. Luykx, D. H. Mathalon, K. A. Mather, V. S. Mattay, S. Matthews, J. Mayoral Van Son, S. C. McEwen, I. Melle, D. W. Morris, B. A. Mueller, M. Nauck, J. E. Nordvik, M. M. Nöthen, D. S. O'Leary, N. Opel, M.-L. P. Martinot, G. B. Pike, A. Preda, E. B. Quinlan, P. E. Rasser, V. Ratnakar, S. Reppermund, V. M. Steen, P. A. Tooney, F. R. Torres, D. J. Veltman, J. T. Voyvodic, R. Whelan, T. White, H. Yamamori, H. H. H. Adams, J. C. Bis, S. Debette, C. Decarli, M. Fornage, V. Gudnason, E. Hofer, M. A. Ikram, L. Launer, W. T. Longstreth, O. L. Lopez, B. Mazoyer, T. H. Mosley, G. V. Roshchupkin, C. L. Satizabal, R. Schmidt, S. Seshadri, Q. Yang, Alzheimer's Disease

Neuroimaging Initiative, CHARGE Consortium, EPIGEN Consortium, IMAGEN Consortium, SYS Consortium, Parkinson's Progression Markers Initiative, M. K. M. Alvim, D. Ames, T. J. Anderson, O. A. Andreassen, A. Arias-Vasquez, M. E. Bastin, B. T. Baune, J. C. Beckham, J. Blangero, D. I. Boomsma, H. Brodaty, H. G. Brunner, R. L. Buckner, J. K. Buitelaar, J. R. Bustillo, W. Cahn, M. J. Cairns, V. Calhoun, V. J. Carr, X. Caseras, S. Caspers, G. L. Cavalleri, F. Cendes, A. Corvin, B. Crespo-Facorro, J. C. Dalrymple-Alford, U. Dannlowski, E. J. C. de Geus, I. J. Deary, N. Delanty, C. Depondt, S. Desrivières, G. Donohoe, T. Espeseth, G. Fernández, S. E. Fisher, H. Flor, A. J. Forstner, C. Francks, B. Franke, D. C. Glahn, R. L. Gollub, H. J. Grabe, O. Gruber, A. K. Håberg, A. R. Hariri, C. A. Hartman, R. Hashimoto, A. Heinz, F. A. Henskens, M. H. J. Hillegers, P. J. Hoekstra, A. J. Holmes, L. E. Hong, W. D. Hopkins, H. E. Hulshoff Pol, T. L. Jernigan, E. G. Jönsson, R. S. Kahn, M. A. Kennedy, T. T. J. Kircher, P. Kochunov, J. B. J. Kwok, S. Le Hellard, C. M. Loughland, N. G. Martin, J.-L. Martinot, C. McDonald, K. L. McMahon, A. Meyer-Lindenberg, P. T. Michie, R. A. Morey, B. Mowry, L. Nyberg, J. Oosterlaan, R. A. Ophoff, C. Pantelis, T. Paus, Z. Pausova, B. W. J. H. Penninx, T. J. C. Polderman, D. Posthuma, M. Rietschel, J. L. Roffman, L. M. Rowland, P. S. Sachdev, P. G. Sämann, U. Schall, G. Schumann, R. J. Scott, K. Sim, S. M. Sisodiya, J. W. Smoller, I. E. Sommer, B. St Pourcain, D. J. Stein, A. W. Toga, J. N. Trollor, N. J. A. Van der Wee, D. van 't Ent, H. Völzke, H. Walter, B. Weber, D. R. Weinberger, M. J. Wright, J. Zhou, J. L. Stein, P. M. Thompson, S. E. Medland, Enhancing NeuroImaging Genetics through Meta-Analysis Consortium (ENIGMA)—Genetics working group, The genetic architecture of the human cerebral cortex, *Science* **367** (2020), doi:10.1126/science.aay6690.

23. J. Z. Liu, S. van Sommeren, H. Huang, S. C. Ng, R. Alberts, A. Takahashi, S. Ripke, J. C. Lee, L. Jostins, T. Shah, S. Abedian, J. H. Cheon, J. Cho, N. E. Dayani, L. Franke, Y. Fuyuno, A. Hart, R. C. Juyal, G. Juyal, W. H. Kim, A. P. Morris, H. Poustchi, W. G. Newman, V. Midha, T. R. Orchard, H. Vahedi, A. Sood, J. Y. Sung, R. Malekzadeh, H.-J. Westra, K. Yamazaki, S.-K. Yang, International Multiple Sclerosis Genetics Consortium, International IBD Genetics Consortium, J. C. Barrett, B. Z. Alizadeh, M. Parkes, T. Bk, M. J. Daly, M. Kubo, C. A. Anderson, R. K. Weersma, Association analyses identify 38 susceptibility loci for inflammatory bowel disease and highlight shared genetic risk across populations, *Nat. Genet.* **47**, 979–986 (2015).

24. C. J. Willer, E. M. Schmidt, S. Sengupta, G. M. Peloso, S. Gustafsson, S. Kanoni, A. Ganna, J. Chen, M. L. Buchkovich, S. Mora, J. S. Beckmann, J. L. Bragg-Gresham, H.-Y. Chang, A. Demirkan, H. M. Den Hertog, R. Do, L. A. Donnelly, G. B. Ehret, T. Esko, M. F. Feitosa, T. Ferreira, K. Fischer, P. Fontanillas, R. M. Fraser, D. F. Freitag, D. Gurdasani, K. Heikkilä, E. Hyppönen, A. Isaacs, A. U. Jackson, Å. Johansson, T. Johnson, M. Kaakinen, J. Kettunen, M. E. Kleber, X. Li, J. 'an Luan, L.-P. Lyytikäinen, P. K. E. Magnusson, M. Mangino, E. Mihailov, M. E. Montasser, M. Müller-Nurasyid, I. M. Nolte, J. R. O'Connell, C. D. Palmer, M. Perola, A.-K. Petersen, S. Sanna, R. Saxena, S. K. Service, S. Shah, D. Shungin, C. Sidore, C. Song, R. J. Strawbridge, I. Surakka, T. Tanaka, T. M. Teslovich, G. Thorleifsson, E. G. Van den Herik, B. F. Voight, K. A. Volcik, L. L. Waite, A. Wong, Y. Wu, W. Zhang, D. Absher, G. Asiki, I. Barroso, L. F. Been, J. L. Bolton, L. L. Bonnycastle, P. Brambilla, M. S. Burnett, G. Cesana, M. Dimitriou, A. S. F. Doney, A. Döring, P. Elliott, S. E. Epstein, G. Ingi Eyjolfsson, B. Gigante, M. O. Goodarzi, H. Grallert, M. L. Gravito, C. J. Groves, G. Hallmans, A.-L. Hartikainen, C. Hayward, D. Hernandez, A. A. Hicks, H. Holm, Y.-J. Hung, T. Illig, M. R. Jones, P. Kaleebu, J. J. P. Kastelein, K.-T. Khaw, E. Kim, N. Klopp, P. Komulainen, M. Kumari, C. Langenberg, T. Lehtimäki, S.-Y. Lin, J. Lindström, R. J. F. Loos, F. Mach, W. L. McArdle, C. Meisinger, B. D. Mitchell, G. Müller, R. Nagaraja, N. Narisu, T. V. M. Nieminen, R. N. Nsubuga, I. Olafsson, K. K. Ong, A. Palotie, T. Papamarkou, C. Pomilla, A. Pouta, D. J. Rader, M. P. Reilly, P. M. Ridker, F. Rivadeneira, I. Rudan, A. Ruokonen, N. Samani, H. Scharnagl, J. Seeley, K. Silander, A. Stančáková, K. Stirrups, A. J. Swift, L. Tiret, A. G. Uitterlinden, L. J. van Pelt, S. Vedantam, N. Wainwright, C. Wijmenga, S. H. Wild, G. Willemsen, T. Wilsgaard, J. F. Wilson, E. H. Young, J. H. Zhao, L. S. Adair, D. Arveiler, T. L. Assimes, S. Bandinelli, F. Bennett, M. Bochud, B. O. Boehm, D. I. Boomsma, I. B. Borecki, S. R. Bornstein, P. Bovet, M. Burnier, H. Campbell, A. Chakravarti, J. C. Chambers, Y.-D. I. Chen, F. S. Collins, R. S. Cooper, J. Danesh, G. Dedoussis, U. de Faire, A. B. Feranil, J. Ferrières, L. Ferrucci, N. B. Freimer, C. Gieger, L. C. Groop, V. Gudnason, U. Gyllensten, A. Hamsten, T. B. Harris, A. Hingorani, J. N. Hirschhorn, A. Hofman, G. K. Hovingh,

C. A. Hsiung, S. E. Humphries, S. C. Hunt, K. Hveem, C. Iribarren, M.-R. Järvelin, A. Jula, M. Kähönen, J. Kaprio, A. Kesäniemi, M. Kivimaki, J. S. Kooner, P. J. Koudstaal, R. M. Krauss, D. Kuh, J. Kuusisto, K. O. Kyvik, M. Laakso, T. A. Lakka, L. Lind, C. M. Lindgren, N. G. Martin, W. März, M. I. McCarthy, C. A. McKenzie, P. Meneton, A. Metspalu, L. Moilanen, A. D. Morris, P. B. Munroe, I. Njølstad, N. L. Pedersen, C. Power, P. P. Pramstaller, J. F. Price, B. M. Psaty, T. Quertermous, R. Rauramaa, D. Saleheen, V. Salomaa, D. K. Sanghera, J. Saramies, P. E. H. Schwarz, W. H.-H. Sheu, A. R. Shuldiner, A. Siegbahn, T. D. Spector, K. Stefansson, D. P. Strachan, B. O. Tayo, E. Tremoli, J. Tuomilehto, M. Uusitupa, C. M. van Duijn, P. Vollenweider, L. Wallentin, N. J. Wareham, J. B. Whitfield, B. H. R. Wolffenbuttel, J. M. Ordovas, E. Boerwinkle, C. N. A. Palmer, U. Thorsteinsdottir, D. I. Chasman, J. I. Rotter, P. W. Franks, S. Ripatti, L. A. Cupples, M. S. Sandhu, S. S. Rich, M. Boehnke, P. Deloukas, S. Kathiresan, K. L. Mohlke, E. Ingelsson, G. R. Abecasis, Global Lipids Genetics Consortium, Discovery and refinement of loci associated with lipid levels, *Nat. Genet.* **45**, 1274–1283 (2013).

25. M. Wuttke, Y. Li, M. Li, K. B. Sieber, M. F. Feitosa, M. Gorski, A. Tin, L. Wang, A. Y. Chu, A. Hoppmann, H. Kirsten, A. Giri, J.-F. Chai, G. Sveinbjornsson, B. O. Tayo, T. Nutile, C. Fuchsberger, J. Marten, M. Cocca, S. Ghasemi, Y. Xu, K. Horn, D. Noce, P. J. van der Most, S. Sedaghat, Z. Yu, M. Akiyama, S. Afaq, T. S. Ahluwalia, P. Almgren, N. Amin, J. Ärnlöv, S. J. L. Bakker, N. Bansal, D. Baptista, S. Bergmann, M. L. Biggs, G. Biino, M. Boehnke, E. Boerwinkle, M. Boissel, E. P. Bottinger, T. S. Boutin, H. Brenner, M. Brumat, R. Burkhardt, A. S. Butterworth, E. Campana, A. Campbell, H. Campbell, M. Canouil, R. J. Carroll, E. Catamo, J. C. Chambers, M.-L. Chee, M.-L. Chee, X. Chen, C.-Y. Cheng, Y. Cheng, K. Christensen, R. Cifkova, M. Ciullo, M. P. Concas, J. P. Cook, J. Coresh, T. Corre, C. F. Sala, D. Cusi, J. Danesh, E. W. Daw, M. H. de Borst, A. De Grandi, R. de Mutsert, A. P. J. de Vries, F. Degenhardt, G. Delgado, A. Demirkan, E. Di Angelantonio, K. Dittrich, J. Divers, R. Dorajoo, K.-U. Eckardt, G. Ehret, P. Elliott, K. Endlich, M. K. Evans, J. F. Felix, V. H. X. Foo, O. H. Franco, A. Franke, B. I. Freedman, S. Freitag-Wolf, Y. Friedlander, P. Froguel, R. T. Gansevoort, H. Gao, P. Gasparini, J. M. Gaziano, V. Giedraitis, C. Gieger, G. Girotto, F. Giulianini, M. Gögele, S. D. Gordon, D. F. Gudbjartsson, V. Gudnason, T. Haller, P. Hamet, T. B. Harris, C. A. Hartman, C. Hayward, J. N. Hellwege, C.-K. Heng, A. A. Hicks, E. Hofer, W. Huang, N. Hutri-Kähönen, S.-J. Hwang, M. A. Ikram, O. S. Indridason, E. Ingelsson, M. Ising, V. W. V. Jaddoe, J. Jakobsdottir, J. B. Jonas, P. K. Joshi, N. S. Josyula, B. Jung, M. Kähönen, Y. Kamatani, C. M. Kammerer, M. Kanai, M. Kastarinen, S. M. Kerr, C.-C. Khor, W. Kiess, M. E. Kleber, W. Koenig, J. S. Kooner, A. Körner, P. Kovacs, A. T. Kraja, A. Krajcoviechova, H. Kramer, B. K. Krämer, F. Kronenberg, M. Kubo, B. Kühnel, M. Kuokkanen, J. Kuusisto, M. La Bianca, M. Laakso, L. A. Lange, C. D. Langefeld, J. J.-M. Lee, B. Lehne, T. Lehtimäki, W. Lieb, Lifelines Cohort Study, S.-C. Lim, L. Lind, C. M. Lindgren, J. Liu, J. Liu, M. Loeffler, R. J. F. Loos, S. Lucae, M. A. Lukas, L.-P. Lyytikäinen, R. Mägi, P. K. E. Magnusson, A. Mahajan, N. G. Martin, J. Martins, W. März, D. Mascalzoni, K. Matsuda, C. Meisinger, T. Meitinger, O. Melander, A. Metspalu, E. K. Mikaelsdottir, Y. Milaneschi, K. Miliku, P. P. Mishra, V. A. Million Veteran Program, K. L. Mohlke, N. Mononen, G. W. Montgomery, D. O. Mook-Kanamori, J. C. Mychaleckyj, G. N. Nadkarni, M. A. Nalls, M. Nauck, K. Nikus, B. Ning, I. M. Nolte, R. Noordam, J. O'Connell, M. L. O'Donoghue, I. Olafsson, A. J. Oldehinkel, M. Orho-Melander, W. H. Ouwehand, S. Padmanabhan, N. D. Palmer, R. Palsson, B. W. J. H. Penninx, T. Perls, M. Perola, M. Pirastu, N. Pirastu, G. Pistis, A. I. Podgornaia, O. Polasek, B. Ponte, D. J. Porteous, T. Poulain, P. P. Pramstaller, M. H. Preuss, B. P. Prins, M. A. Province, T. J. Rabelink, L. M. Raffield, O. T. Raitakari, D. F. Reilly, R. Rettig, M. Rheinberger, K. M. Rice, P. M. Ridker, F. Rivadeneira, F. Rizzi, D. J. Roberts, A. Robino, P. Rossing, I. Rudan, R. Rueedi, D. Ruggiero, K. A. Ryan, Y. Saba, C. Sabanayagam, V. Salomaa, E. Salvi, K.-U. Saum, H. Schmidt, R. Schmidt, B. Schöttker, C.-A. Schulz, N. Schupf, C. M. Shaffer, Y. Shi, A. V. Smith, B. H. Smith, N. Soranzo, C. N. Spracklen, K. Strauch, H. M. Stringham, M. Stumvoll, P. O. Svensson, S. Szymczak, E.-S. Tai, S. M. Tajuddin, N. Y. Q. Tan, K. D. Taylor, A. Teren, Y.-C. Tham, J. Thiery, C. H. L. Thio, H. Thomsen, G. Thorleifsson, D. Toniolo, A. Tönjes, J. Tremblay, I. Tzoulaki, A. G. Uitterlinden, S. Vaccargiu, R. M. van Dam, P. van der Harst, C. M. van Duijn, D. R. Velez Edward, N. Verweij, S. Vogelezang, U. Völker, P. Vollenweider, G. Waeber, M. Waldenberger, L. Wallentin, Y. X. Wang, C. Wang, D. M. Waterworth, W. Bin Wei, H. White, J. B. Whitfield, S. H. Wild, J. F. Wilson, M. K. Wojczynski, C. Wong, T.-Y. Wong, L. Xu, Q. Yang, M. Yasuda, L. M. Yerges-Armstrong, W.

Zhang, A. B. Zonderman, J. I. Rotter, M. Bochud, B. M. Psaty, V. Vitart, J. G. Wilson, A. Dehghan, A. Parsa, D. I. Chasman, K. Ho, A. P. Morris, O. Devuyst, S. Akilesh, S. A. Pendergrass, X. Sim, C. A. Böger, Y. Okada, T. L. Edwards, H. Snieder, K. Stefansson, A. M. Hung, I. M. Heid, M. Scholz, A. Teumer, A. Köttgen, C. Pattaro, A catalog of genetic loci associated with kidney function from analyses of a million individuals, *Nat. Genet.* **51**, 957–972 (2019).

32. e1071.pdf, (available at https://cran.r-project.org/web/packages/e1071/e1071.pdf).

33. P. H. Lee, V. Anttila, H. Won, Y.-C. A. Feng, J. Rosenthal, Z. Zhu, E. M. Tucker-Drob, M. G. Nivard, A. D. Grotzinger, D. Posthuma, M. M. -J. Wang, D. Yu, E. A. Stahl, R. K. Walters, R. J. L. Anney, L. E. Duncan, T. Ge, R. Adolfsson, T. Banaschewski, S. Belangero, E. H. Cook, G. Coppola, E. M. Derks, P. J. Hoekstra, J. Kaprio, A. Keski-Rahkonen, G. Kirov, H. R. Kranzler, J. J. Luykx, L. A. Rohde, C. C. Zai, E. Agerbo, M. J. Arranz, P. Asherson, M. Bækvad-Hansen, G. Baldursson, M. Bellgrove, R. A. Belliveau, J. Buitelaar, C. L. Burton, J. Bybjerg-Grauholm, M. Casas, F. Cerrato, K. Chambert, C. Churchhouse, B. Cormand, J. Crosbie, S. Dalsgaard, D. Demontis, A. E. Doyle, A. Dumont, J. Elia, J. Grove, O. O. Gudmundsson, J. Haavik, H. Hakonarson, C. S. Hansen, C. A. Hartman, Z. Hawi, A. Hervás, D. M. Hougaard, D. P. Howrigan, H. Huang, J. Kuntsi, K. Langley, K.-P. Lesch, P. W. L. Leung, S. K. Loo, J. Martin, A. R. Martin, J. J. McGough, S. E. Medland, J. L. Moran, O. Mors, P. B. Mortensen, R. D. Oades, D. S. Palmer, C. B. Pedersen, M. G. Pedersen, T. Peters, T. Poterba, J. B. Poulsen, J. A. Ramos-Quiroga, A. Reif, M. Ribasés, A. Rothenberger, P. Rovira, C. Sánchez-Mora, F. Kyle Satterstrom, R. Schachar, M. S. Artigas, S. Steinberg, H. Stefansson, P. Turley, G. Bragi Walters, T. Werge, T. Zayats, D. E. Arking, F. Bettella, J. D. Buxbaum, J. H. Christensen, R. L. Collins, H. Coon, S. De Rubeis, R. Delorme, D. E. Grice, T. F. Hansen, P. A. Holmans, S. Hope, C. M. Hultman, L. Klei, C. Ladd-Acosta, P. Magnusson, T. Nærland, M. Nyegaard, D. Pinto, P. Qvist, K. Rehnström, A. Reichenberg, J. Reichert, K. Roeder, G. A. Rouleau, E. Saemundsen, S. J. Sanders, S. Sandin, B. St Pourcain, K. Stefansson, J. S. Sutcliffe, M. E. Talkowski, L. A. Weiss, A. Jeremy Willsey, I. Agartz, H. Akil, D. Albani, M. Alda, T. D. Als, A. Anjorin, L. Backlund, N. Bass, M. Bauer, B. T. Baune, F. Bellivier, S. E. Bergen, W. H. Berrettini, J. M. Biernacka, D. H. R. Blackwood, E. Bøen, M. Budde, W. Bunney, M. Burmeister, W. Byerley, E. M. Byrne, S. Cichon, T.-K. Clarke, J. R. I. Coleman, N. Craddock, D. Curtis, P. M. Czerski, A. M. Dale, N. Dalkner, U. Dannlowski, F. Degenhardt, A. Di Florio, T. Elvsåshagen, B. Etain, S. B. Fischer, A. J. Forstner, L. Forty, J. Frank, M. Frye, J. M. Fullerton, K. Gade, H. A. Gaspar, E. S.

Gershon, M. Gill, F. S. Goes, S. D. Gordon, K. Gordon-Smith, M. J. Green, T. A. Greenwood, M. Grigoroiu-Serbanescu, J. Guzman-Parra, J. Hauser, M. Hautzinger, U. Heilbronner, S. Herms, P. Hoffmann, D. Holland, S. Jamain, I. Jones, L. A. Jones, R. Kandaswamy, J. R. Kelsoe, J. L. Kennedy, O. K. Joachim, S. Kittel-Schneider, M. Kogevinas, A. C. Koller, C. Lavebratt, C. M. Lewis, Q. S. Li, J. Lissowska, L. M. O. Loohuis, S. Lucae, A. Maaser, U. F. Malt, N. G. Martin, L. Martinsson, S. L. McElroy, F. J. McMahon, A. McQuillin, I. Melle, A. Metspalu, V. Millischer, P. B. Mitchell, G. W. Montgomery, G. Morken, D. W. Morris, B. Müller-Myhsok, N. Mullins, R. M. Myers, C. M. Nievergelt, M. Nordentoft, A. N. Adolfsson, M. M. Nöthen, R. A. Ophoff, M. J. Owen, S. A. Paciga, C. N. Pato, M. T. Pato, R. H. Perlis, A. Perry, J. B. Potash, C. S. Reinbold, M. Rietschel, M. Rivera, M. Roberson, M. Schalling, P. R. Schofield, T. G. Schulze, L. J. Scott, A. Serretti, E. Sigurdsson, O. B. Smeland, E. Stordal, F. Streit, J. Strohmaier, T. E. Thorgeirsson, J. Treutlein, G. Turecki, A. E. Vaaler, E. Vieta, J. B. Vincent, Y. Wang, S. H. Witt, P. Zandi, R. A. H. Adan, L. Alfredsson, T. Ando, H. Aschauer, J. H. Baker, V. Bencko, A. W. Bergen, A. Birgegård, V. B. Perica, H. Brandt, R. Burghardt, L. Carlberg, M. Cassina, M. Clementi, P. Courtet, S. Crawford, S. Crow, J. J. Crowley, U. N. Danner, O. S. P. Davis, D. Degortes, J. E. DeSocio, D. M. Dick, C. Dina, E. Docampo, K. Egberts, S. Ehrlich, T. Espeseth, F. Fernández-Aranda, M. M. Fichter, L. Foretova, M. Forzan, G. Gambaro, I. Giegling, F. Gonidakis, P. Gorwood, M. G. Mayora, Y. Guo, K. A. Halmi, K. Hatzikotoulas, J. Hebebrand, S. G. Helder, B. Herpertz-Dahlmann, W. Herzog, A. Hinney, H. Imgart, S. Jiménez-Murcia, C. Johnson, J. Jordan, A. Julià, D. Kaminská, L. Karhunen, A. Karwautz, M. J. H. Kas, W. H. Kaye, M. A. Kennedy, Y.-R. Kim, L. Klareskog, K. L. Klump, G. P. S. Knudsen, M. Landén, S. Le Hellard, R. D. Levitan, D. Li, P. Lichtenstein, M. Maj, S. Marsal, S. McDevitt, J. Mitchell, P. Monteleone, A. M. Monteleone, M. A. Munn-Chernoff, B. Nacmias, M. Navratilova, J. K. O’Toole, L. Padyukov, J. Pantel, H. Papezova, R. Rabionet, A. Raevuori, N. Ramoz, T. Reichborn-Kjennerud, V. Ricca, M. Roberts, D. Rujescu, F. Rybakowski, A. Scherag, U. Schmidt, J. Seitz, L. Slachtova, M. C. Slof-Op‘t, A. Slopien, S. Sorbi, L. Southam, M. Strober, A. Tortorella, F. Tozzi, J. Treasure, K. Tziouvas, A. A. van Elburg, T. D. Wade, G. Wagner, E. Walton, H. J. Watson, H.-E. Wichmann, D. Blake Woodside, E. Zeggini, S. Zerwas, S. Zipfel, M. J. Adams, T. F. M. Andlauer, K. Berger, E. B. Binder, D. I. Boomsma, E. Castelao, L. Colodro-Conde, N. Direk, A. R. Docherty, E. Domenici, K. Domschke, E. C. Dunn, J. C. Foo, E. J. de., H. J. Grabe, S. P. Hamilton, C. Horn, J.-J. Hottenga, D. Howard, M. Ising, S. Kloiber, D. F. Levinson, G. Lewis, P. K. E. Magnusson, H. Mbarek, C. M. Middeldorp, S. Mostafavi, D. R. Nyholt, B. W. Penninx, R. E. Peterson, G. Pistis, D. J. Porteous, M. Preisig, J. A. Quiroz, C. Schaefer, E. C. Schulte, J. Shi, D. J. Smith, P. A. Thomson, H. Tiemeier, R. Uher, S. van der Auwera, M. M. Weissman, M. Alexander, M. Begemann, E. Bramon, N. G. Buccola, M. J. Cairns, D. Campion, V. J. Carr, C. Robert Cloninger, D. Cohen, D. A. Collier, A. Corvin, L. E. DeLisi, G. Donohoe, F. Dudbridge, J. Duan, R. Freedman, P. V. Gejman, V. Golimbet, S. Godard, H. Ehrenreich, A. M. Hartmann, F. A. Henskens, M. Ikeda, N. Iwata, A. V. Jablensky, I. Joa, E. G. Jönsson, B. J. Kelly, J. Knight, B. Konte, C. Laurent-Levinson, J. Lee, T. Lencz, B. Lerer, C. M. Loughland, A. K. Malhotra, J. Mallet, C. McDonald, M. Mitjans, B. J. Mowry, K. C. Murphy, R. M. Murray, F. Anthony O’Neill, S.-Y. Oh, A. Palotie, C. Pantelis, A. E. Pulver, T. L. Petryshen, D. J. Quested, B. Riley, A. R. Sanders, U. Schall, S. G. Schwab, R. J. Scott, P. C. Sham, J. M. Silverman, K. Sim, A. A. Steixner, P. A. Tooney, J. van Os, M. P. Vawter, D. Walsh, M. Weiser, D. B. Wildenauer, N. M. Williams, B. K. Wormley, F. Zhang, C. Androutsos, P. D. Arnold, C. L. Barr, C. Barta, K. Bey, O. Joseph Bienvenu, D. W. Black, L. W. Brown, C. Budman, D. Cath, K.-A. Cheon, V. Ciullo, B. J. Coffey, D. Cusi, L. K. Davis, D. Denys, C. Depienne, A. Dietrich, V. Eapen, P. Falkai, T. V. Fernandez, B. Garcia-Delgar, D. A. Geller, D. L. Gilbert, M. A. Grados, E. Greenberg, E. Grünblatt, J. Hagstrøm, G. L. Hanna, A. Hartmann, T. Hedderly, G. A. Heiman, I. Heyman, H. J. Hong, A. Huang, C. Huyser, L. Ibanez-Gomez, E. A. Khramtsova, Y. K. Kim, Y.-S. Kim, R. A. King, Y.-J. Koh, A. Konstantinidis, S. Kook, S. Kuperman, B. L. Leventhal, C. Lochner, A. G. Ludolph, M. Madruga-Garrido, I. Malaty, A. Maras, J. T. McCracken, I. A. Meijer, P. Mir, A. Morer, K. R. Müller-Vahl, A. Münchau, T. L. Murphy, A. Naarden, P. Nagy, G. Nestadt, P. S. Nestadt, H. Nicolini, E. L. Nurmi, M. S. Okun, P. Paschou, F. Piras, F. Piras, C. Pittenger, K. J. Plessen, M. A. Richter, R. Rizzo, M. Robertson, V. Roessner, S. Ruhrmann, J. F. Samuels, P. Sandor, M. Schlögelhofer, E.-Y. Shin, H. Singer, D.-H. Song, J. Song, G. Spalletta, D. J. Stein, S. Evelyn Stewart, E. A. Storch, B. Stranger, M. Stuhrmann, Z. Tarnok, J. A. Tischfield, J. Tübing, F. Visscher, N. Vulink, M. Wagner, S. Walitza, S. Wanderer, M. Woods, Y. Worbe, G. Zai, S. H. Zinner, P. F. Sullivan, B. Franke, M. J.

Daly, C. M. Bulik, A. M. McIntosh, M. C. O'Donovan, A. Zheutlin, O. A. Andreassen, A. D. Børglum, G. Breen, H. J. Edenberg, A. H. Fanous, S. V. Faraone, J. Gelernter, C. A. Mathews, M. Mattheisen, K. S. Mitchell, M. C. Neale, J. I. Nurnberger, S. Ripke, S. L. Santangelo, J. M. Scharf, M. B. Stein, L. M. Thornton, J. T. R. Walters, N. R. Wray, D. H. Geschwind, B. M. Neale, K. S. Kendler, J. W. Smoller, Genomic Relationships, Novel Loci, and Pleiotropic Mechanisms across Eight Psychiatric Disorders, *Cell* **179**, 1469–1482.e11 (2019).

34. Cross-Disorder Group of the Psychiatric Genomics Consortium, S. H. Lee, S. Ripke, B. M. Neale, S. V. Faraone, S. M. Purcell, R. H. Perlis, B. J. Mowry, A. Thapar, M. E. Goddard, J. S. Witte, D. Absher, I. Agartz, H. Akil, F. Amin, O. A. Andreassen, A. Anjorin, R. Anney, V. Anttila, D. E. Arking, P. Asherson, M. H. Azevedo, L. Backlund, J. A. Badner, A. J. Bailey, T. Banaschewski, J. D. Barchas, M. R. Barnes, T. B. Barrett, N. Bass, A. Battaglia, M. Bauer, M. Bayés, F. Bellivier, S. E. Bergen, W. Berrettini, C. Betancur, T. Bettecken, J. Biederman, E. B. Binder, D. W. Black, D. H. R. Blackwood, C. S. Bloss, M. Boehnke, D. I. Boomsma, G. Breen, R. Breuer, R. Bruggeman, P. Cormican, N. G. Buccola, J. K. Buitelaar, W. E. Bunney, J. D. Buxbaum, W. F. Byerley, E. M. Byrne, S. Caesar, W. Cahn, R. M. Cantor, M. Casas, A. Chakravarti, K. Chambert, K. Choudhury, S. Cichon, C. R. Cloninger, D. A. Collier, E. H. Cook, H. Coon, B. Cormand, A. Corvin, W. H. Coryell, D. W. Craig, I. W. Craig, J. Crosbie, M. L. Cuccaro, D. Curtis, D. Czamara, S. Datta, G. Dawson, R. Day, E. J. De Geus, F. Degenhardt, S. Djurovic, G. J. Donohoe, A. E. Doyle, J. Duan, F. Dudbridge, E. Duketis, R. P. Ebstein, H. J. Edenberg, J. Elia, S. Ennis, B. Etain, A. Fanous, A. E. Farmer, I. N. Ferrier, M. Flickinger, E. Fombonne, T. Foroud, J. Frank, B. Franke, C. Fraser, R. Freedman, N. B. Freimer, C. M. Freitag, M. Friedl, L. Frisén, L. Gallagher, P. V. Gejman, L. Georgieva, E. S. Gershon, D. H. Geschwind, I. Giegling, M. Gill, S. D. Gordon, K. Gordon-Smith, E. K. Green, T. A. Greenwood, D. E. Grice, M. Gross, D. Grozeva, W. Guan, H. Gurling, L. De Haan, J. L. Haines, H. Hakonarson, J. Hallmayer, S. P. Hamilton, M. L. Hamshere, T. F. Hansen, A. M. Hartmann, M. Hautzinger, A. C. Heath, A. K. Henders, S. Herms, I. B. Hickie, M. Hipolito, S. Hoefels, P. A. Holmans, F. Holsboer, W. J. Hoogendijk, J.-J. Hottenga, C. M. Hultman, V. Hus, A. Ingason, M. Ising, S. Jamain, E. G. Jones, I. Jones, L. Jones, J.-Y. Tzeng, A. K. Kähler, R. S. Kahn, R. Kandaswamy, M. C. Keller, J. L. Kennedy, E. Kenny, L. Kent, Y. Kim, G. K. Kirov, S. M. Klauck, L. Klei, J. A. Knowles, M. A. Kohli, D. L. Koller, B. Konte, A. Korszun, L. Krabbendam, R. Krasucki, J. Kuntsi, P. Kwan, M. Landén, N. Långström, M. Lathrop, J. Lawrence, W. B. Lawson, M. Leboyer, D. H. Ledbetter, P. H. Lee, T. Lencz, K.-P. Lesch, D. F. Levinson, C. M. Lewis, J. Li, P. Lichtenstein, J. A. Lieberman, D.-Y. Lin, D. H. Linszen, C. Liu, F. W. Lohoff, S. K. Loo, C. Lord, J. K. Lowe, S. Lucae, D. J. MacIntyre, P. A. F. Madden, E. Maestrini, P. K. E. Magnusson, P. B. Mahon, W. Maier, A. K. Malhotra, S. M. Mane, C. L. Martin, N. G. Martin, M. Mattheisen, K. Matthews, M. Mattingsdal, S. A. McCarroll, K. A. McGhee, J. J. McGough, P. J. McGrath, P. McGuffin, M. G. McInnis, A. McIntosh, R. McKinney, A. W. McLean, F. J. McMahon, W. M. McMahon, A. McQuillin, H. Medeiros, S. E. Medland, S. Meier, I. Melle, F. Meng, J. Meyer, C. M. Middeldorp, L. Middleton, V. Milanova, A. Miranda, A. P. Monaco, G. W. Montgomery, J. L. Moran, D. Moreno-De-Luca, G. Morken, D. W. Morris, E. M. Morrow, V. Moskvina, P. Muglia, T. W. Mühleisen, W. J. Muir, B. Müller-Myhsok, M. Murtha, R. M. Myers, I. Myin-Germeys, M. C. Neale, S. F. Nelson, C. M. Nievergelt, I. Nikolov, V. Nimgaonkar, W. A. Nolen, M. M. Nöthen, J. I. Nurnberger, E. A. Nwulia, D. R. Nyholt, C. O'Dushlaine, R. D. Oades, A. Olincy, G. Oliveira, L. Olsen, R. A. Ophoff, U. Osby, M. J. Owen, A. Palotie, J. R. Parr, A. D. Paterson, C. N. Pato, M. T. Pato, B. W. Penninx, M. L. Pergadia, M. A. Pericak-Vance, B. S. Pickard, J. Pimm, J. Piven, D. Posthuma, J. B. Potash, F. Poustka, P. Propping, V. Puri, D. J. Quested, E. M. Quinn, J. A. Ramos-Quiroga, H. B. Rasmussen, S. Raychaudhuri, K. Rehnström, A. Reif, M. Ribasés, J. P. Rice, M. Rietschel, K. Roeder, H. Roeyers, L. Rossin, A. Rothenberger, G. Rouleau, D. Ruderfer, D. Rujescu, A. R. Sanders, S. J. Sanders, S. L. Santangelo, J. A. Sergeant, R. Schachar, M. Schalling, A. F. Schatzberg, W. A. Scheftner, G. D. Schellenberg, S. W. Scherer, N. J. Schork, T. G. Schulze, J. Schumacher, M. Schwarz, E. Scolnick, L. J. Scott, J. Shi, P. D. Shilling, S. I. Shyn, J. M. Silverman, S. L. Slager, S. L. Smalley, J. H. Smit, E. N. Smith, E. J. S. Sonuga-Barke, D. St Clair, M. State, M. Steffens, H.-C. Steinhausen, J. S. Strauss, J. Strohmaier, T. S. Stroup, J. S. Sutcliffe, P. Szatmari, S. Szelinger, S. Thirumalai, R. C. Thompson, A. A. Todorov, F. Tozzi, J. Treutlein, M. Uhr, E. J. C. G. van den Oord, G. Van Grootheest, J. Van Os, A. M. Vicente, V. J. Vieland, J. B. Vincent, P. M. Visscher, C. A. Walsh, T. H. Wassink, S. J.

Watson, M. M. Weissman, T. Werge, T. F. Wienker, E. M. Wijsman, G. Willemsen, N. Williams, A. J. Willsey, S. H. Witt, W. Xu, A. H. Young, T. W. Yu, S. Zammit, P. P. Zandi, P. Zhang, F. G. Zitman, S. Zöllner, B. Devlin, J. R. Kelsoe, P. Sklar, M. J. Daly, M. C. O'Donovan, N. Craddock, P. F. Sullivan, J. W. Smoller, K. S. Kendler, N. R. Wray, International Inflammatory Bowel Disease Genetics Consortium (IIBDGC), Genetic relationship between five psychiatric disorders estimated from genome-wide SNPs, *Nat. Genet.* **45**, 984–994 (2013).

35. M. Nagel, P. R. Jansen, S. Stringer, K. Watanabe, C. A. de Leeuw, J. Bryois, J. E. Savage, A. R. Hammerschlag, N. G. Skene, A. B. Muñoz-Manchado, 23andMe Research Team, T. White, H. Tiemeier, S. Linnarsson, J. Hjerling-Leffler, T. J. C. Polderman, P. F. Sullivan, S. van der Sluis, D. Posthuma, Meta-analysis of genome-wide association studies for neuroticism in 449,484 individuals identifies novel genetic loci and pathways, *Nat. Genet.* **50**, 920–927 (2018).

36. M. J. Gandal, J. R. Haney, N. N. Parikshak, V. Leppa, G. Ramaswami, C. Hartl, A. J. Schork, V. Appadurai, A. Buil, T. M. Werge, C. Liu, K. P. White, CommonMind Consortium, PsychENCODE Consortium, iPSYCH-BROAD Working Group, S. Horvath, D. H. Geschwind, Shared molecular neuropathology across major psychiatric disorders parallels polygenic overlap, *Science* **359**, 693–697 (2018).

37. S. Sniekers, S. Stringer, K. Watanabe, P. R. Jansen, J. R. I. Coleman, E. Krapohl, E. Taskesen, A. R. Hammerschlag, A. Okbay, D. Zabaneh, N. Amin, G. Breen, D. Cesarini, C. F. Chabris, W. G. Iacono, M. A. Ikram, M. Johannesson, P. Koellinger, J. J. Lee, P. K. E. Magnusson, M. McGue, M. B. Miller, W. E. R. Ollier, A. Payton, N. Pendleton, R. Plomin, C. A. Rietveld, H. Tiemeier, C. M. van Duijn, D. Posthuma, Genome-wide association meta-analysis of 78,308 individuals identifies new loci and genes influencing human intelligence, *Nat. Genet.* **49**, 1107–1112 (2017).
